# Environmental risk factors associated with the presence of *Mycobacterium ulcerans* in Victoria, Australia

**DOI:** 10.1101/2022.01.10.22269030

**Authors:** Kim R. Blasdell, Bridgette McNamara, Daniel P. O’Brien, Mary Tachedjian, Victoria Boyd, Michael Dunn, Peter T. Mee, Simone Clayton, Julie Gaburro, Ina Smith, Katherine B. Gibney, Ee Laine Tay, Emma C Hobbs, Nilakshi Waidyatillake, Stacey E. Lynch, Timothy P. Stinear, Eugene Athan

## Abstract

In recent years reported cases of Buruli ulcer (BU), caused by *Mycobacterium ulcerans* (MU), have increased substantially in Victoria, Australia, with the epidemic also expanding geographically. To develop an understanding of how MU circulates in the environment and transmits to humans we analyzed environmental samples collected from 115 properties of recent BU cases and from 115 postcode-matched control properties, for the presence of MU. Environmental factors associated with increased odds of MU presence at a property included certain native plant species and native vegetation in general, more alkaline soil, lower altitude, the presence of common ringtail possums (*Pseudocheirus peregrinus*) and overhead powerlines. However, only powerlines and the absence of the native plant *Melaleuca lanceolata* were associated with BU case properties. Samples positive for MU were more likely to be found at case properties and were associated with detections of MU in ringtail possum feces, supporting the hypothesis that MU is zoonotic, with ringtail possums the strongest reservoir host candidate. However, the disparity in environmental risk factors associated with MU positive properties versus case properties indicates a strong human behavioral component or the influence of other environmental factors in disease acquisition that requires further study.

**Article Summary Line:** Possums, powerlines, and native vegetation are associated with the presence of *Mycobacterium ulcerans* in residential properties in Victoria, Australia.

## Introduction

Buruli ulcer (BU) is a neglected tropical disease, caused by the environmental pathogen *Mycobacterium ulcerans* (MU). Affecting all age groups, the disease causes severe destructive lesions of skin and soft tissue and results in significant morbidity, sometimes leading to long term disability and deformity (O’Brien et al., 2015). Endemic to more than 30 countries, the highest disease burden is in sub-Saharan Africa (Johnson et al., 2005; O’Brien et al., 2019). Case numbers have increased in Australia (O’Brien et al., 2018; O’Brien et al., 2019), most markedly in the temperate, southern state of Victoria (Loftus et al., 2018) where case numbers increased from 32 in 2010 to a peak of 340 in 2018, with 217 cases in 2020 and 208 cases reported up to October 2021 (Victorian DH, 2021a). The endemic area is also expanding geographically, with new disease hotspots reported both in Victoria’s second largest city (Geelong; Victorian DH, 2019) and most recently in Melbourne’s inner suburbs (Victorian DH, 2021b; Tai et al., 2018).

Previous studies have identified several risk factors and potential transmission routes. In Africa, BU foci are often associated with natural water bodies (Aiga et al., 2004; Bratschi et al., 2014) and in Victoria, an outbreak was linked to exposure to a contaminated water irrigation system at a golf course (Veitch et al., 1997). In a questionnaire-based case control study in one Victorian hotspot, the risk of having BU was found to be increased in people who did not wash minor skin wounds immediately, did not frequently wear insect repellent or long trousers outdoors, and who received mosquito bites to the lower legs or arms (Quek et al., 2007). Molecular detection of MU in mosquitoes collected from several localities within the Victorian endemic area (Johnson et al., 2007; Lavender et al., 2011), and the demonstration that *Ae. notoscriptus* can act as mechanical vectors for BU in a mouse model (Wallace et al., 2017) suggests that mosquitoes may be involved with BU transmission in Victoria. Several studies have also suggested that MU may be a zoonotic pathogen in Victoria. Evidence of infection and disease has been reported in several native and non-native mammals (Elsner et al., 2008; Mitchell et al., 1987; O’Brien et al., 2011; O’Brien et al., 2013; O’Brien et al., 2014; Van Zyl et al., 2010), but there is increasing evidence that two common possum species may be acting as reservoirs hosts in south east Australia (Fyfe et al., 2010). Both common brushtail (BT; *Trichosurus vulpecula*) and common ringtail (RT; *Pseudocheirus peregrinus*) possums can develop BU and possum feces are the environmental sample type most commonly PCR positive for MU in Victorian endemic areas (Fyfe et al., 2010; O’Brien et al., 2014). There is evidence of a clear geographic correlation between the presence of human cases and MU*-*positive possum feces (Carson et al., 2014; Fyfe et al., 2010).

Here we present the environmental results from the first systematic, large-scale case-control study to encompass almost the entire Victorian endemic area. By assessing the environmental characteristics of participants’ gardens and the distribution of MU in different environmental sample types within these, we establish: (1) which environmental sources are more predictive of MU presence (including predicting the presence of viable bacteria); (2) what features make a property more likely to be positive for MU or more likely to contain a human case of BU; and (3) how MU status changes through time. These findings will aid public education around this disease and inform the development of intervention strategies to prevent disease.

## Methods

### Ethics

The study was approved by the Victorian Department of Health (DH) Human Research Ethics Committee and the CSIRO Health and Medical Human Research Ethics Committee (application no. 10/18). Access to electoral information for medical research purposes was granted by the Australian Electoral Commission. Written informed consent was obtained for the property environmental field surveys.

### Study area

The study was conducted in the known Buruli ulcer-endemic area of Victoria, Australia. This is primarily located around Port Phillip Bay, with the main concentration of recognized cases from the Mornington and Bellarine Peninsulas and the Melbourne regional (Bayside) area.

### Recruitment

All laboratory confirmed BU cases (Betts at al, 2018) aged ≥18 years notified to the Victorian DH between 12^th^ June 2018 and 11^th^ June 2020 were eligible to participate. Potential control participants (aged ≥18 years) were randomly selected from either the 2017 Victorian Population Health Survey (VPHS) or the Australian Electoral Roll. Participants were asked to complete a paper-based questionnaire (results to be reported elsewhere). Environmental surveys were conducted on a subset of case and control properties within the endemic area.

***Case properties*** had at least one resident with a laboratory-confirmed diagnosis of BU within the study period (12^th^ June 2018 to 11^th^ June 2020). ***Control properties*** had no residents diagnosed with BU within the study period or reported as having had BU prior to the study period.

Cases who completed the study questionnaire were purposely selected by postcode to ensure a representative spread of sampling across the affected area based on reported BU prevalence (i.e. more properties were surveyed in postcodes with more cases). Control properties were then purposely selected based on postcode and matched 1:1 to case properties.

### Property environmental field surveys

Prior to an environmental field survey being conducted at a property, geocoordinates (latitude and longitude), altitude (all from https://www.google.com/maps) and approximate property size (https://www.freemaptools.com/area-calculator.htm) were recorded and an outline of the property, including buildings was prepared. During the property visit, additional information was recorded, including presence of key plant species (Supplemental Figure 1), garden type and samples collected (Supplemental Figure 2; Supplemental Figure 3, Supplemental material: Field Collection Sheet). Garden type was categorized as Non-native (>60% non-native vegetation), Mixed (40-60% native/non-native) or Native (>60% native vegetation). Five different sample types were collected as outlined in Table 1, namely soil, water, plants (Supplemental Figure 4), feces and insects. Soil texture was determined as per standard protocols (https://www.dpi.nsw.gov.au/ data/assets/pdf_file/0008/168866/texture-salinity.pdf). Up to 20 environmental samples were collected per property. In addition, a mains water sample was also collected from each property as a negative control, to validate sample collection techniques and detect potential contamination. The total number of observable water sources on a property was recorded, although samples were not always collected from all sources. The presence of mosquito larvae in any of the water sources was also recorded.

**Table 1:**
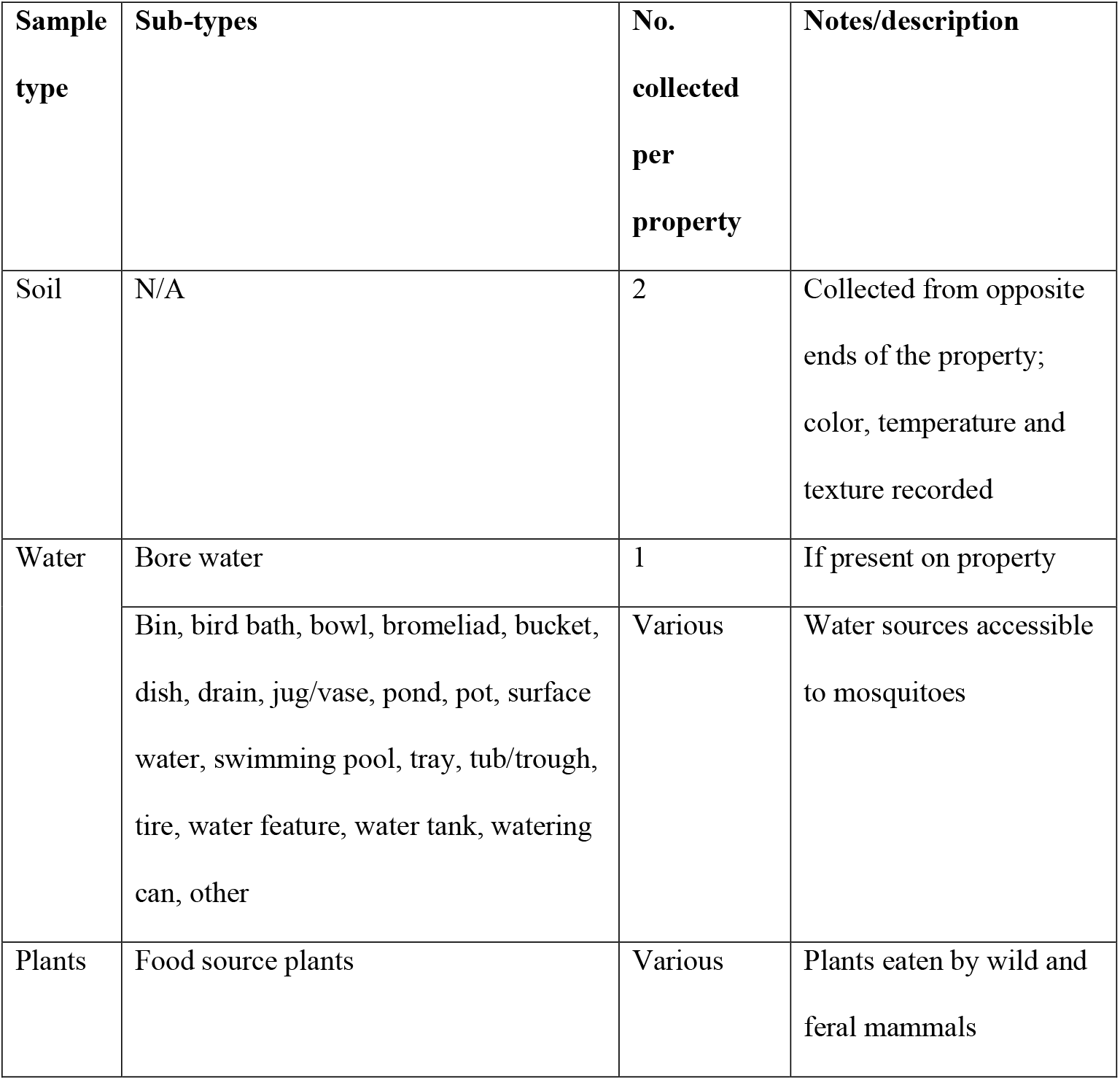
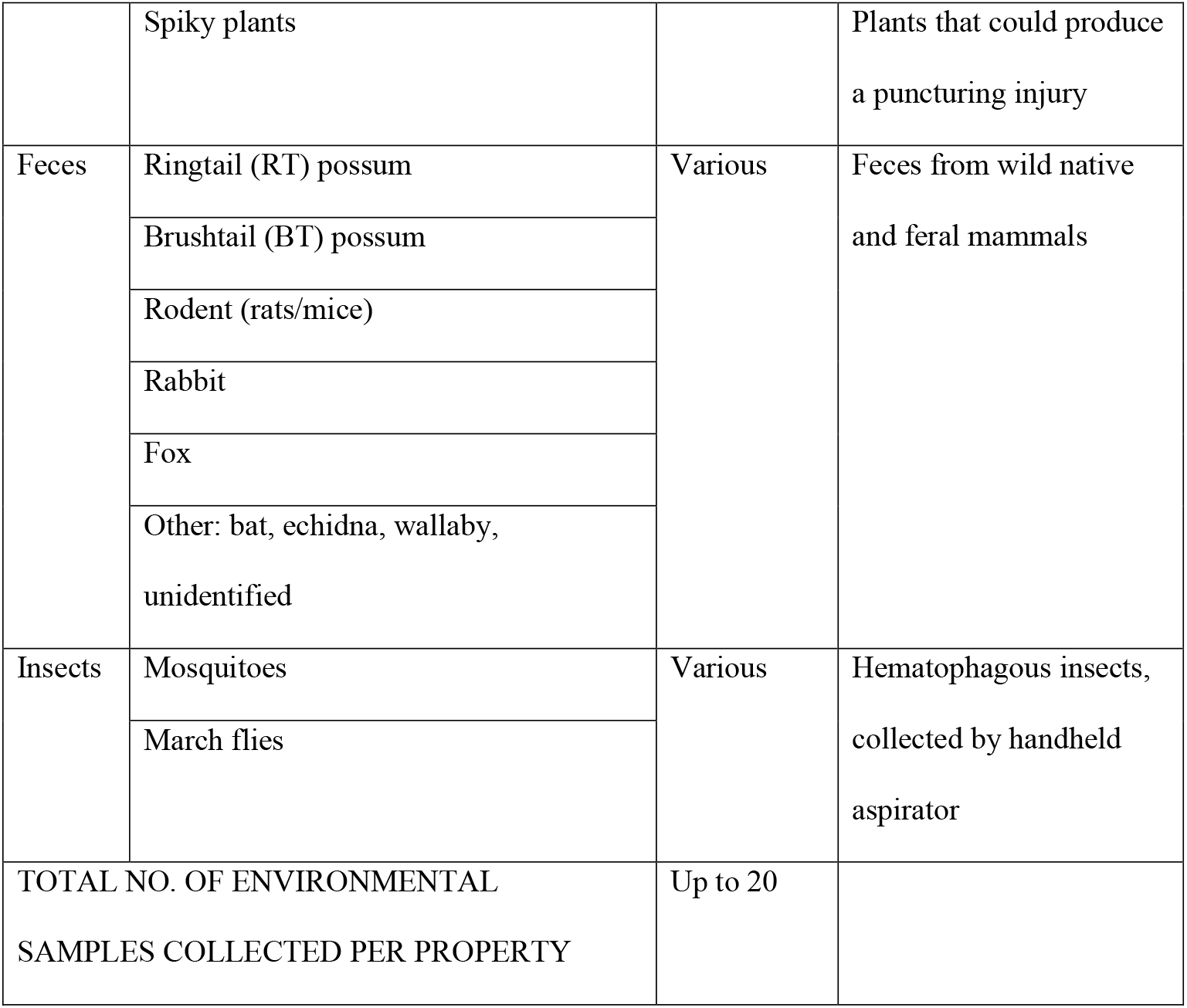
Environmental field survey sample type categories. The mains water negative control is not included in the property sample total.

| **Sample type** | **Sub-types** | **No. collected per property** | **Notes/description** |
| --- | --- | --- | --- |
| Soil | N/A | 2 | Collected from opposite ends of the property; color, temperature and texture recorded |
| Water | Bore water | 1 | If present on property |
|  | Bin, bird bath, bowl, bromeliad, bucket, dish, drain, jug/vase, pond, pot, surface water, swimming pool, tray, tub/trough, tire, water feature, water tank, watering can, other | Various | Water sources accessible to mosquitoes |
| Plants | Food source plants | Various | Plants eaten by wild and feral mammals |
|  | Spiky plants |  | Plants that could produce a puncturing injury |
| Feces | Ringtail (RT) possum | Various | Feces from wild native and feral mammals |
|  | Brushtail (BT) possum |  |  |
|  | Rodent (rats/mice) |  |  |
|  | Rabbit |  |  |
|  | Fox |  |  |
|  | Other: bat, echidna, wallaby, unidentified |  |  |
| Insects | Mosquitoes | Various | Hematophagous insects, collected by handheld aspirator |
|  | March flies |  |  |
| TOTAL NO. OF ENVIRONMENTAL SAMPLES COLLECTED PER PROPERTY |  | Up to 20 |  |

A proportion of properties were visited twice. For these properties first visits were made between 5th August 2019 and 3rd March 2020, whilst return visits were made between 19th March 2020 and 23rd June 2020. The interval between visits was impacted by work and travel restrictions imposed during the COVID-19 pandemic. For all return environmental field surveys at a property, the property outline from the initial visit was used to enable the same sample types to be collected from the same locations. Any different samples collected and significant environmental changes between the two field surveys were recorded. All samples were returned to the laboratory and maintained at -70°C until processed.

### Laboratory processing and analysis

Soil samples were processed to determine soil bulk density (g/cm^3^), pH, conductivity (μS/cm) and salinity class. For soil bulk density, 50cm^3^ of each soil sample was weighed before and after heating in an oven at 105°C for two hours and the dry weight divided by the soil volume. For pH and conductivity, soil was resuspended in distilled water at a 1:5 ratio, before testing with a VisionPlus pH/EC80 meter (Jenco). Soil salinity class was determined based on the meter reading for conductivity with reference to the soil texture type determined during the field survey (https://www.agric.wa.gov.au/soil-salinity/measuring-soil-salinity). Samples for nucleic acid extraction were thawed prior to processing and transferred to 2ml tubes containing approximately 2.4g of a mixture of 2.3mm and 0.5mm zirconia/silica beads (Bio Spec Products, Inc.). Water samples were added in 500μl volumes to 500μl of DNA/RNA Shield (Zymo). For plant, soil and fecal samples, approximately 0.2g (plants) or 0.1g (feces/soil) was added to 1ml of DNA/RNA Shield (Zymo). Samples were homogenized at 6500rpm for 30sec on a Precellys 24 (Bertin Technologies) and clarified for 5 mins at 16,000g. Total nucleic acid was extracted from 200μl of the cleared supernatant using the Kingfisher Flex benchtop automated extraction instrument (ThermoFisher) and the Quick DNA/RNA MagBead Pathogen kit (Zymo) as per the manufacturer’s instructions. All samples were subjected to the IS2404 real-time PCR assay, which is routinely used for the molecular diagnosis of MU infection in clinical samples and has been used previously on environmental samples (Fyfe et al., 2007). As this assay detects other mycolactone-producing *Mycobacteria* in addition to MU, any sample that tested positive by this assay (CT<40, threshold 0.02) was subjected to confirmatory testing using two MU specific assays (IS2606 and KR; Fyfe et al., 2007) as well as an RNA-based assay targeting the MU 16S rRNA to assess viability (Beissner et al., 2012). This viability assay can also detect some strains of *M. marinum*, although it is unlikely that this species would be present in most of the sample types collected. Based on the results of these assays, all samples were classified as negative (IS2404 not detected or CT≥40); IS2404 detected (CT<40, threshold 0.02); confirmed (MU detected by both IS2606 and KR assays); or viable (MU 16S rRNA detected) (Figure 1). A property was assigned an ‘IS2404 not detected’ or ‘negative’ status if IS2404 was not detected in any samples collected from that property and was classified as IS2404 detected / confirmed / viable if any samples collected from that property met these definitions (N.B. a ‘viable’ property would also be both ‘IS2404 detected’ and ‘confirmed’; a ‘confirmed’ property would also be ‘IS2404 detected’).

**Figure 1:**
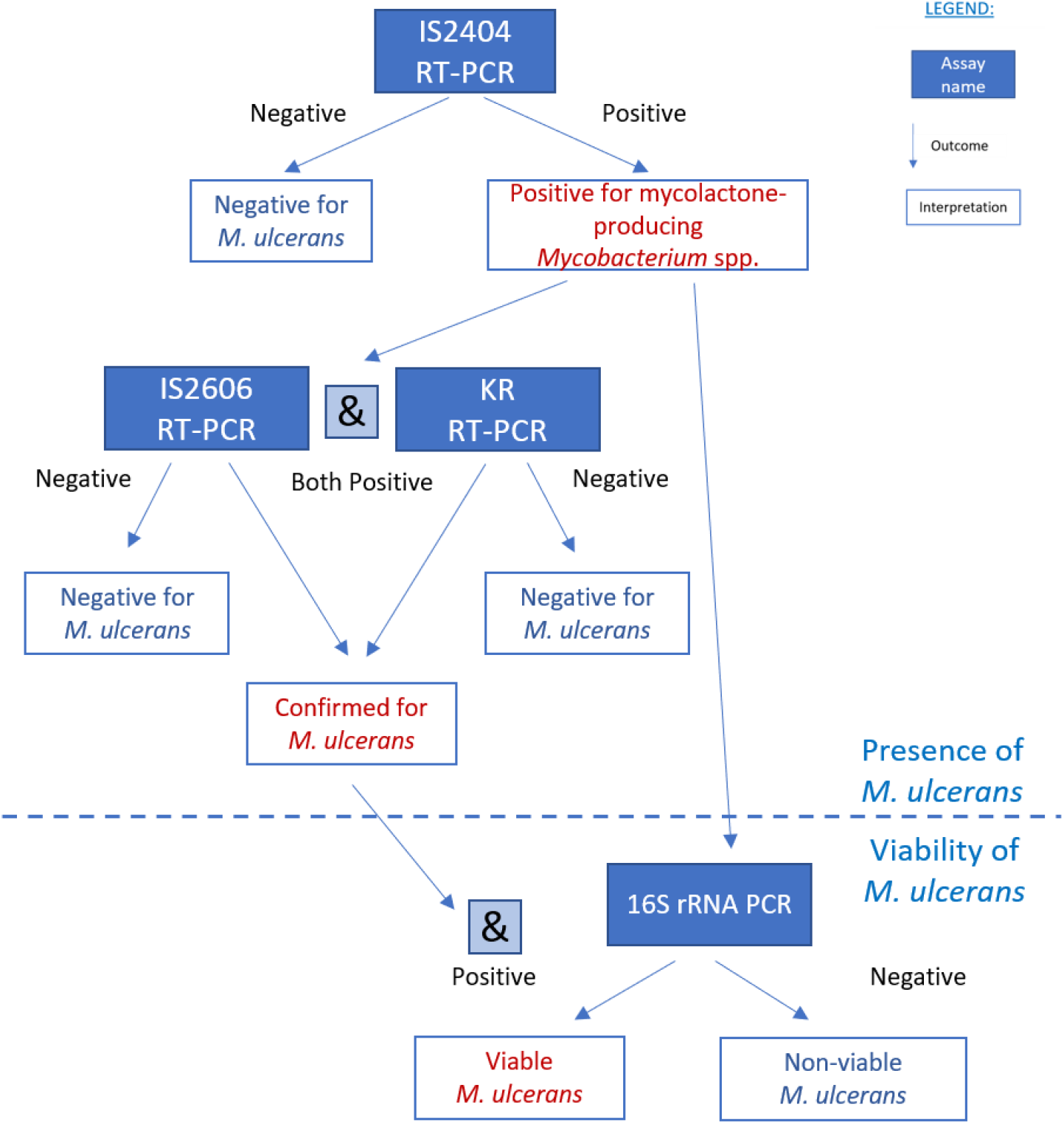
Flow diagram for sample processing and interpretation of results based on the different RT-PCR assay results.

### Statistical analysis

We investigated relationships between each of the property outcomes (IS2404 detected, confirmed, viable and case status) with environmental variables including garden type, and property location, size and altitude. For initial analysis Fisher’s Exact (where expected values ≤5) and chi-square tests were used to compare categorical variables, and either Student’s t-test or one-way ANOVA with a post-hoc Tukey Kramer test were used to compare differences in mean values of continuous variables. Logistic regression models were used to estimate the strength of the relationships between each of the selected environmental characteristics and i) properties with one or more sample positive for MU, or ii) case status of the property, expressed as odds ratios (OR) and 95% confidence intervals (95%CI). The potential for confounding by covariates was assessed using a subject matter informed directed acyclic graph (DAG) and appropriate confounders included in each adjusted model (Supplemental Figure 5), along with variables that had P<0.1 in the univariate analysis and were identified in 30 or more properties. Descriptive statistical tests were conducted using pre-prepared spreadsheets available from http://www.biostathandbook.com (McDonald, 2014). Regression models were conducted using Stata 15 (Statacorp).

## Results

### Numbers sampled: Participants/Field surveys/Samples/Assays

Of the 3,433 individuals contacted, 283/497 (56.9%) cases and 520/2936 (17.7%) controls participated in the case-control study. Of these, 256 (90.5%) case participants and 458 (88.1%) control participants agreed to be contacted about the property environmental surveys. Property environmental field surveys were conducted at 230 properties, comprising 115 case and 115 control properties, located across 20 postcodes (Figure 2). A second field survey was carried out within three to nine months of the initial survey at 27 properties (13 case properties; 14 control properties), all located within three of the most severely impacted postcodes.

A total of 4363 environmental samples (excluding insect samples) were collected during the field surveys, 3907 from initial surveys and 456 from return surveys (Table 2). Of these, 475 (10.9%) samples were ‘IS2404 detected’ (highest for feces (20.5%) and soil (13.2%)) and 237 (5.4%) samples were ‘confirmed’, most commonly for feces in general (13.3%), and for fox and RT possum feces in particular (20.0% and 16.7% respectively)) (Table 2). Of ‘IS2404 detected’ samples, considerably higher proportions of feces were also ‘confirmed’ compared to other sample types. Sixty-seven samples (1.5% of all samples) were ‘viable’. At least one sample from each sample type was IS2404 detected and confirmed, however only feces were ‘viable’ – most frequently from RT possums (64 samples) but also from two BT possums and a fox.

**Figure 2:**
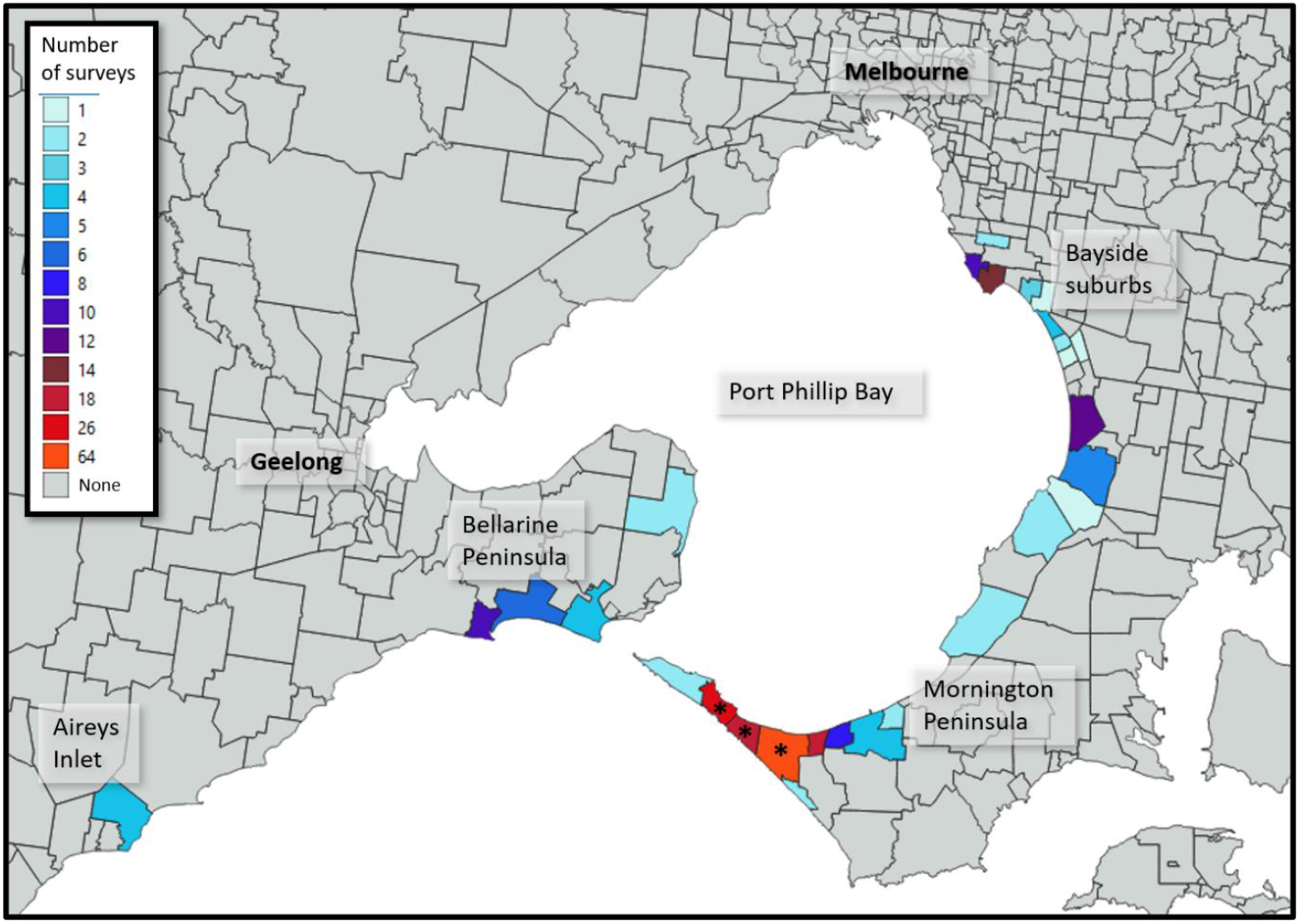
Map of affected area, illustrating the number of property surveys conducted by suburb. An Asterix (*) is shown on suburbs where repeat sampling was undertaken. N.B. Geographical boundaries are not available by postcode and some postcodes contain more than one suburb.

**Table 2:**
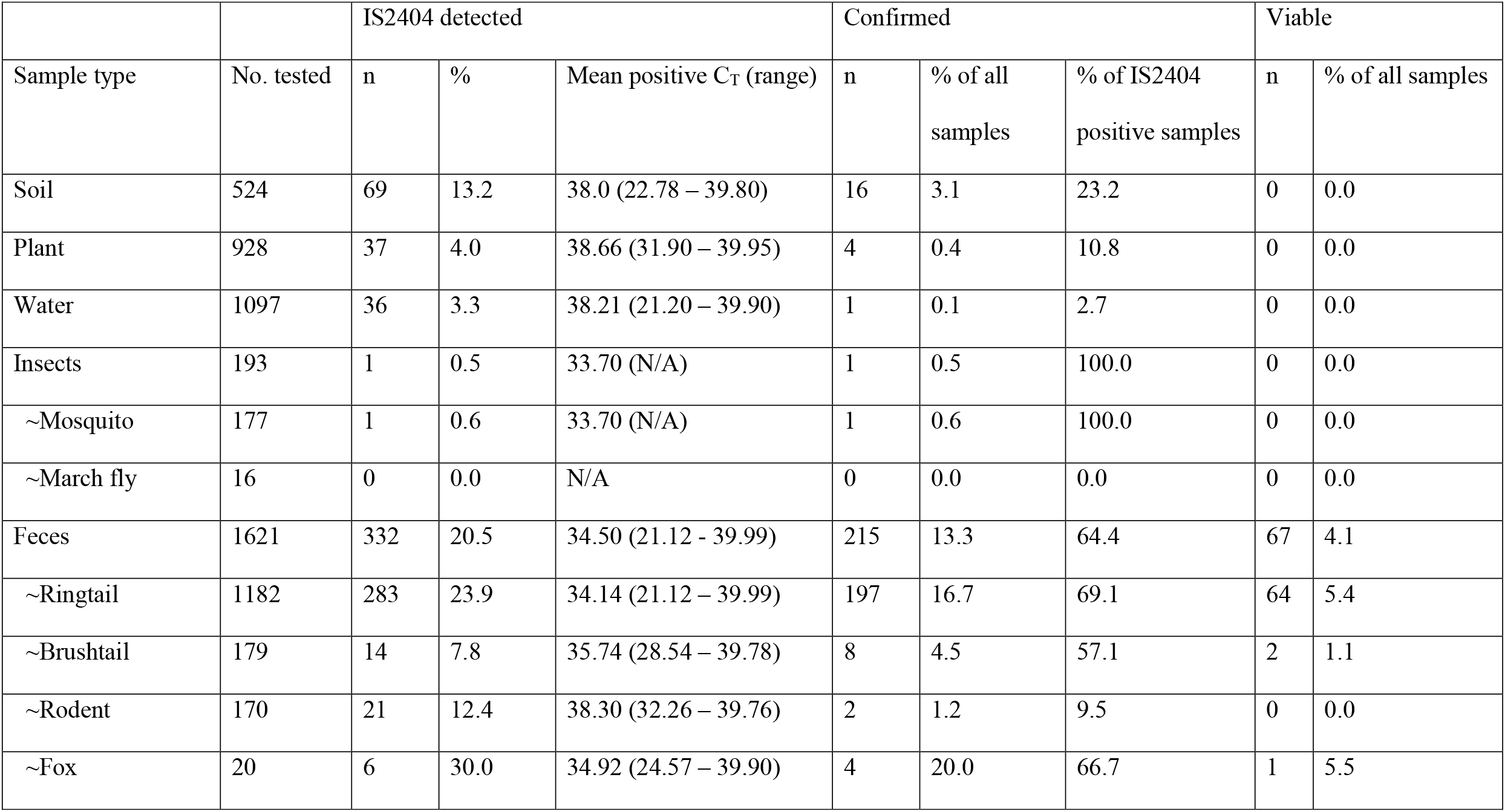
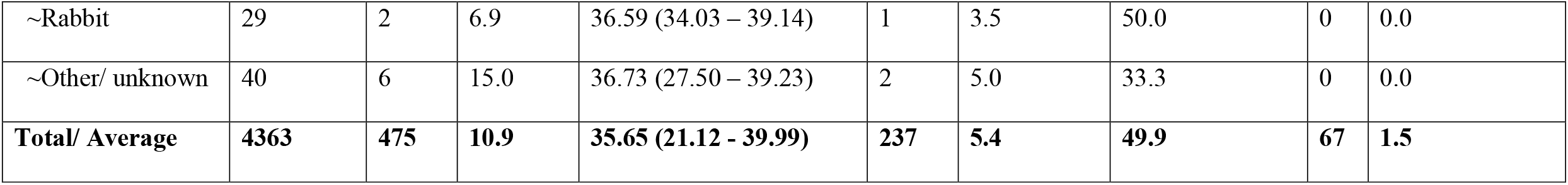
Results of sample testing by sample type, with sub-type shown for fecal and insect samples.

|  |  | IS2404 detected |  |  | Confirmed |  |  | Viable |  |
| --- | --- | --- | --- | --- | --- | --- | --- | --- | --- |
| Sample type | No. tested | n | % | Mean positive C<sub>T</sub> (range) | n | % of all samples | % of IS2404 positive samples | n | % of all samples |
| Soil | 524 | 69 | 13.2 | 38.0 (22.78 – 39.80) | 16 | 3.1 | 23.2 | 0 | 0.0 |
| Plant | 928 | 37 | 4.0 | 38.66 (31.90 – 39.95) | 4 | 0.4 | 10.8 | 0 | 0.0 |
| Water | 1097 | 36 | 3.3 | 38.21 (21.20 – 39.90) | 1 | 0.1 | 2.7 | 0 | 0.0 |
| Insects | 193 | 1 | 0.5 | 33.70 (N/A) | 1 | 0.5 | 100.0 | 0 | 0.0 |
| ~Mosquito | 177 | 1 | 0.6 | 33.70 (N/A) | 1 | 0.6 | 100.0 | 0 | 0.0 |
| ~March fly | 16 | 0 | 0.0 | N/A | 0 | 0.0 | 0.0 | 0 | 0.0 |
| Feces | 1621 | 332 | 20.5 | 34.50 (21.12 - 39.99) | 215 | 13.3 | 64.4 | 67 | 4.1 |
| ~Ringtail | 1182 | 283 | 23.9 | 34.14 (21.12 – 39.99) | 197 | 16.7 | 69.1 | 64 | 5.4 |
| ~Brushtail | 179 | 14 | 7.8 | 35.74 (28.54 – 39.78) | 8 | 4.5 | 57.1 | 2 | 1.1 |
| ~Rodent | 170 | 21 | 12.4 | 38.30 (32.26 – 39.76) | 2 | 1.2 | 9.5 | 0 | 0.0 |
| ~Fox | 20 | 6 | 30.0 | 34.92 (24.57 – 39.90) | 4 | 20.0 | 66.7 | 1 | 5.5 |
| ~Rabbit | 29 | 2 | 6.9 | 36.59 (34.03 – 39.14) | 1 | 3.5 | 50.0 | 0 | 0.0 |
| ~Other/ unknown | 40 | 6 | 15.0 | 36.73 (27.50 – 39.23) | 2 | 5.0 | 33.3 | 0 | 0.0 |
| **Total/ Average** | **4363** | **475** | **10.9** | **35.65 (21.12 - 39.99)** | **237** | **5.4** | **49.9** | **67** | **1.5** |

For the initial surveys, 157/230 (68.3%) properties were IS2404 detected, of which 103 (44.8%) were confirmed and 46 (20.0%) viable. For the second (return) surveys, 16/27 (59.3%) were IS2404 detected, nine (33.3%) confirmed and two (7.4%) viable. Among case properties, the interval between case notification date and field collection date did not affect the odds of a property testing IS2404 detected, confirmed or viable (data on request). At individual properties, a maximum of 10 samples were IS2404 detected, six samples were confirmed, and four samples were viable. Of the 20 postcodes in which properties were surveyed, at least one property was IS2404 detected in 17 postcodes, confirmed in 13 postcodes, and viable in ten postcodes (Supplemental Figure 6).

### Comparison of PCR assays

Excluding insect samples (due to a single positive), IS2404 CT values differed significantly between the sample types (one-way ANOVA, p<0.001), although the ‘between sample type’ variance was considerably lower than the ‘within sample type’ variance (26.7% and 73.3% respectively). Fecal CT values were significantly lower (suggesting higher bacterial loads) (mean = 34.50) than those for all other sample types (mean = 38.23 combined, Tukey-Kramer test, p<0.05). There were no significant differences in CT values between the other sample types (means: plant = 38.66; soil = 38.0; water = 38.21). Lower IS2404 CT values were observed for confirmed samples (median = 33.69; IQR = 7.20) than unconfirmed samples (median = 38.79; IQR = 1.11). Only 8/238 (3.3%) of IS2404-positive samples that were unconfirmed had CT values of <35.

Case properties were more likely to be ‘IS2404 detected’ and ‘confirmed’ than control properties when considering all samples and when restricted to fecal samples or RT possum feces only (Table 3). No significant relationships were observed for the viability assay.

**Table 3:** Significant relationships observed between case properties and sample status as assessed by Chi-square test

| Status | Factor | No. (%) case properties | No. (%) control properties | p-value |
| --- | --- | --- | --- | --- |
| IS2404 detected | Any IS2404 detected sample | 88 (76.5) | 69 (60.0) | 0.007 |
|  | IS2404 detected feces | 73 (63.5) | 50 (43.5) | 0.002 |
|  | IS2404 detected ringtail feces | 63 (54.8) | 46 (40.0) | 0.025 |
| Confirmed | Any confirmed sample | 63 (54.8) | 40 (34.8) | 0.002 |
|  | Confirmed feces | 59 (50.9) | 36 (31.3) | 0.002 |
|  | Confirmed ringtail feces | 54 (47.0) | 34 (29.6) | 0.007 |

### Environmental characteristics of different property types

Mean property size varied between the different study areas (ANOVA, p<0.05). Sampled properties in the Mornington Peninsula (n=148 properties, postcodes 3930-3944, mean property size 1087m^2^) were larger than those in Bayside (n=56, postcodes 3190-3199, mean 695m^2^) but did not differ significantly from sampled properties in Bellarine (n=22, postcodes 3223-3227, mean=890m^2^) or the Surf Coast (n=4, postcode 3231, mean = 677m^2^). No significant differences were found between area and altitude, with average property elevation ranging between 13.5m (Bellarine) to 22.75m (Bayside).

### Univariate analysis of property characteristics and study outcomes (IS2404 detected, confirmed, viable and case status)

Univariate analyses are presented in Table 4 (unadjusted OR) and Supplementary Table 1. Due to the close association between garden type and the presence of selected native plant species, the former was not included in the multivariable models, despite properties with native gardens having higher odds of being IS2404 detected, confirmed and viable. Due to low sample size, the presence of rabbit feces was also not included in the multivariable models, even though properties with rabbit feces were more likely to have viable MU at the property. Two significant relationships that were observed in univariate analyses but not in multivariable analyses were the positive association between *Melaleuca lanceolata* and IS2404 detected, confirmed and viable properties, and the higher soil salinity associated with IS2404 and confirmed properties.

**Table 4:**
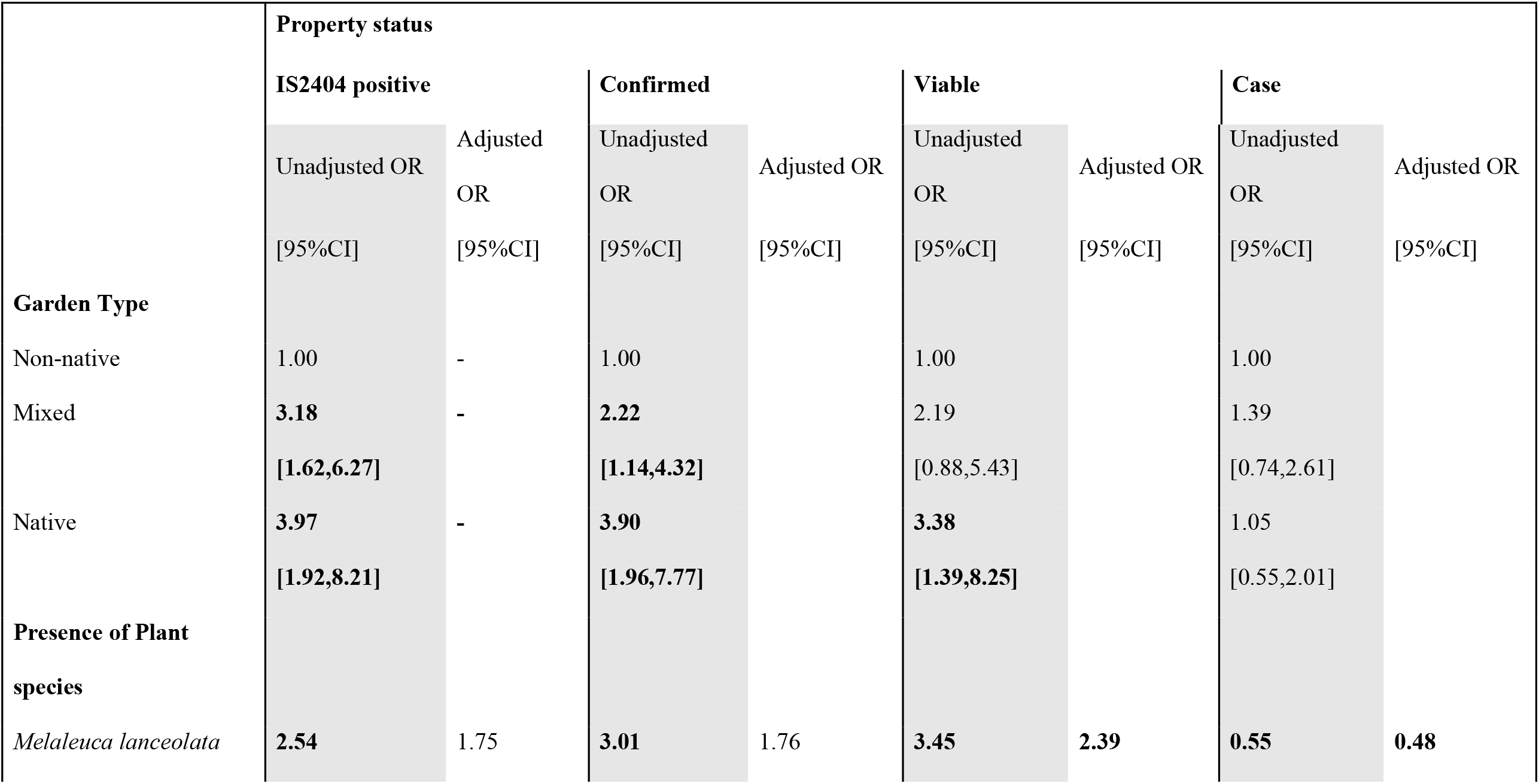
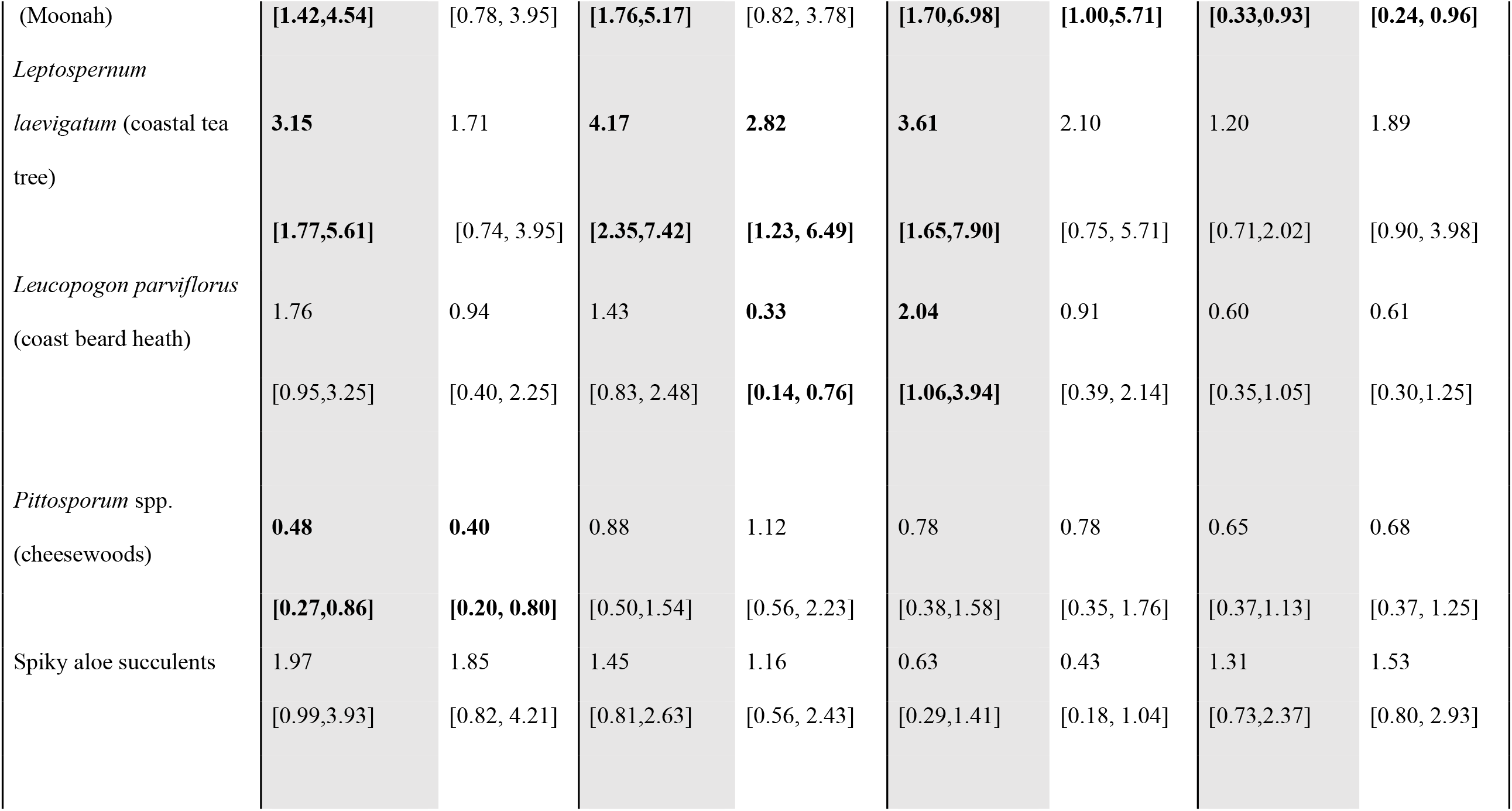
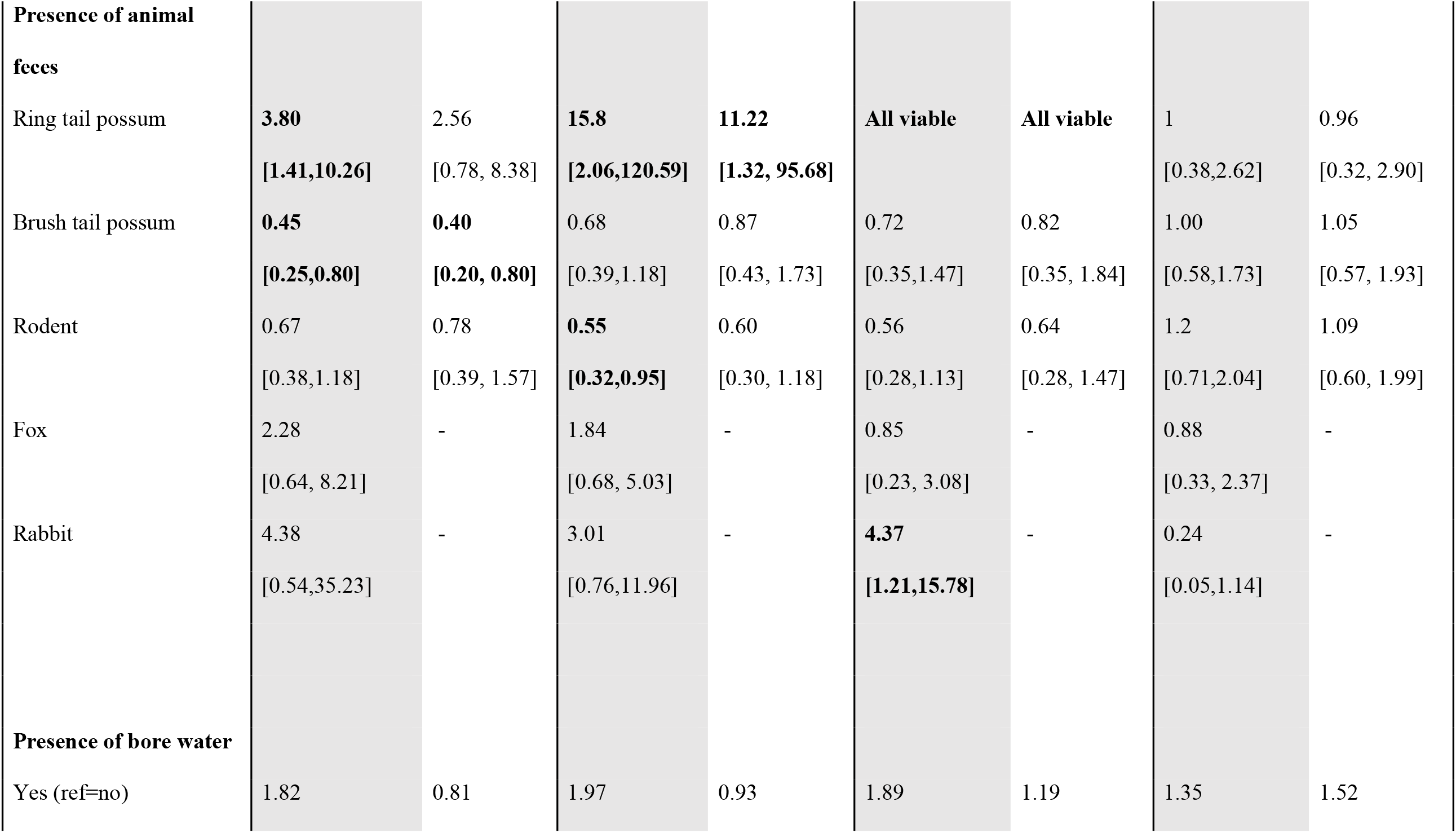
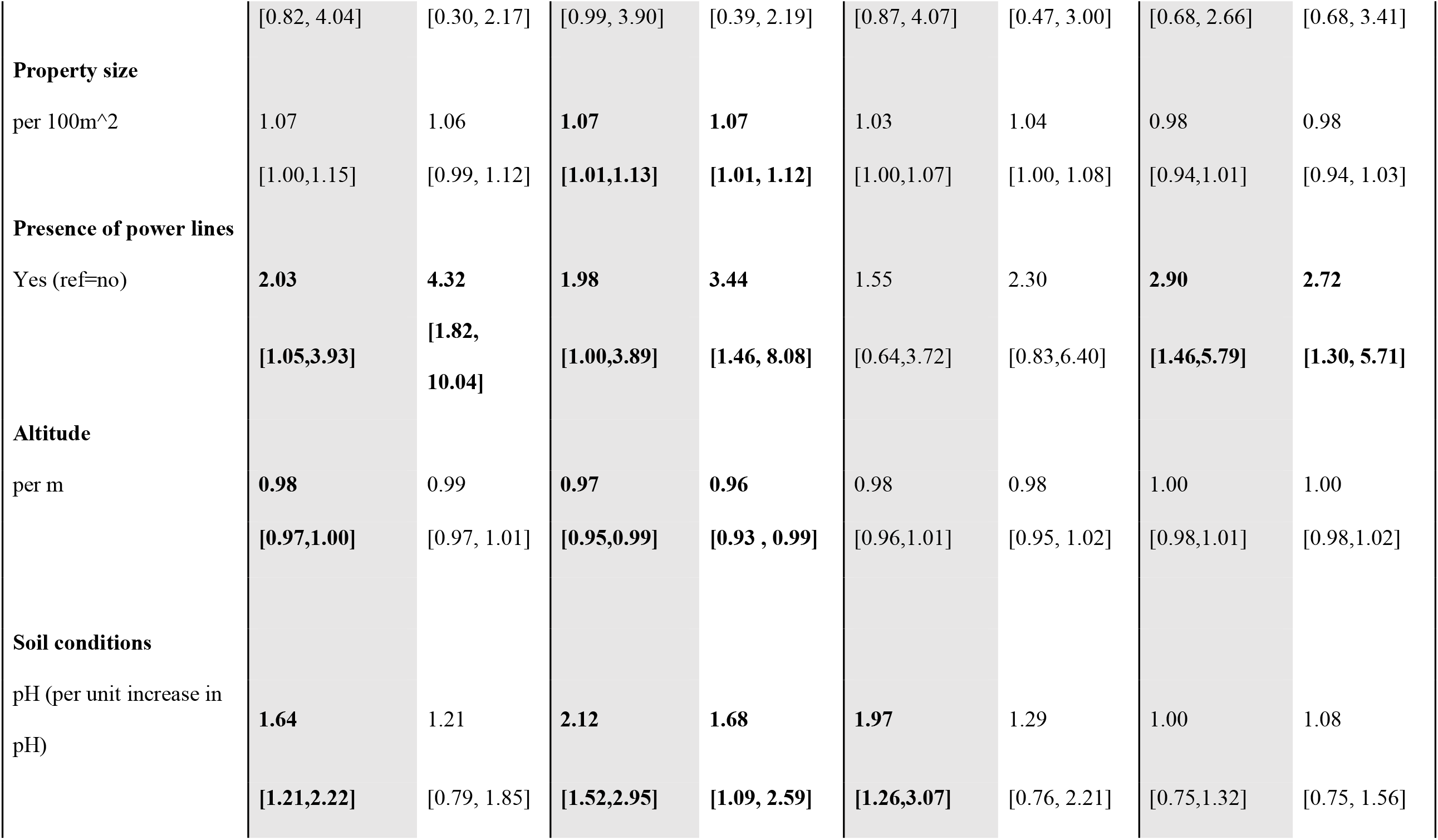
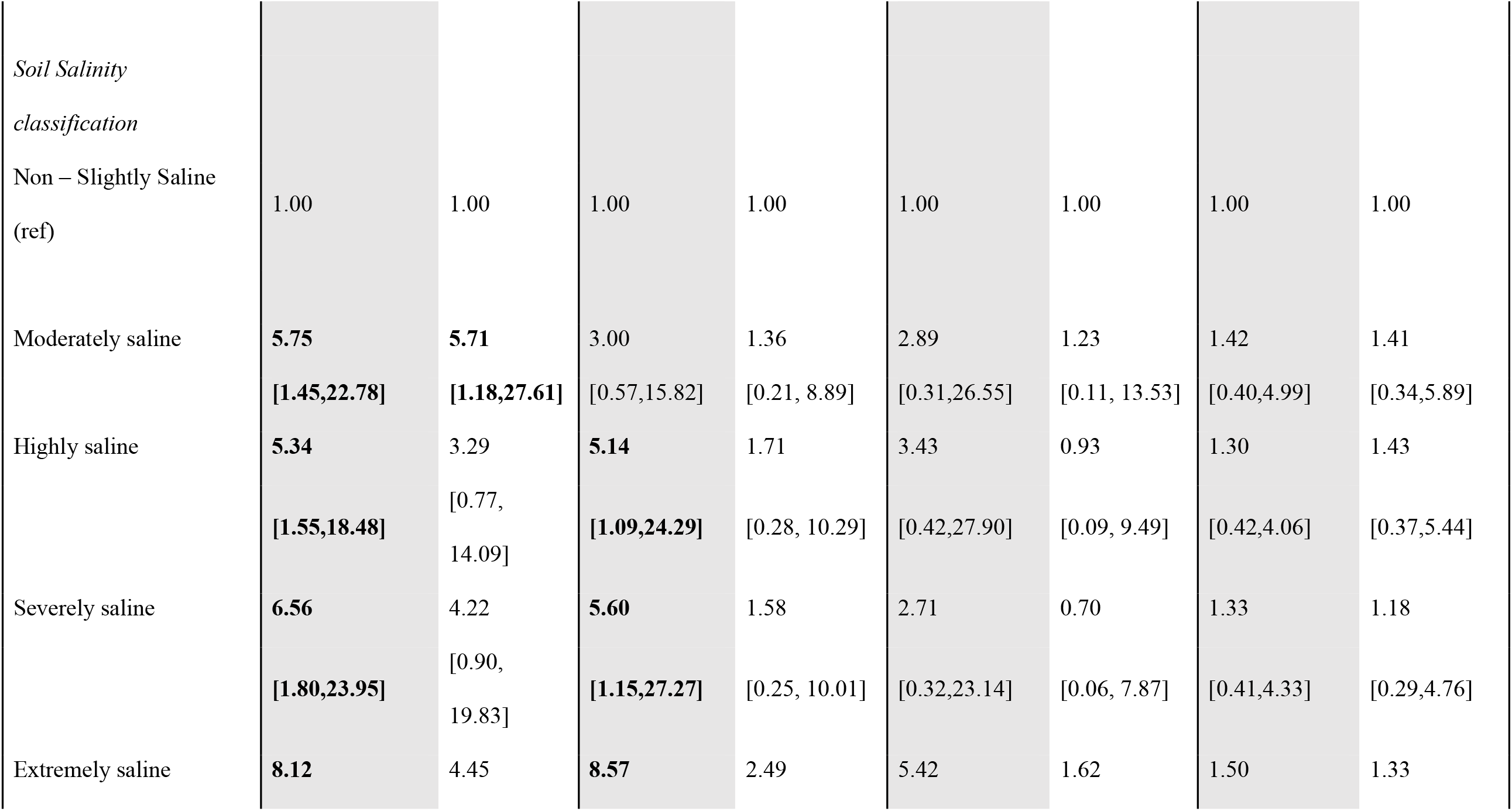
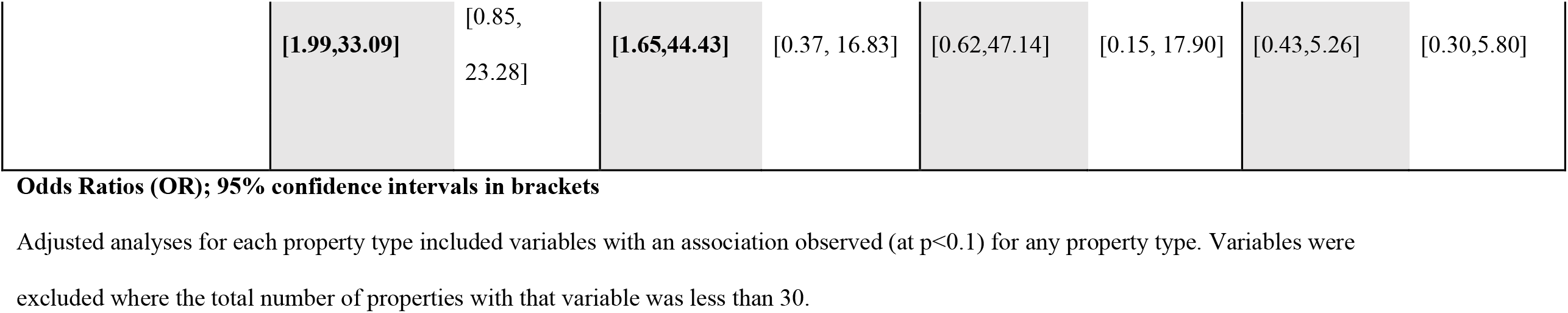
Relationships between environmental characteristics and MU property status (IS2404 positive, confirmed, viable) or case status

### Multivariable analysis of property characteristics and study outcomes (IS2404, confirmed, viable and case status)

In multivariable analysis, the presence of selected plant species was associated with both increased odds of property status (*Leptospernum laevigatum* for confirmed properties; *M. lanceolata* (Moonah) for viable properties) and decreased odds of property status (*M. lanceolata* (Moonah) for case properties; *Leucopogon parviflorus* (coastal beard heath) for confirmed properties; and *Pittosporum* (cheesewoods) for IS2404 properties). Likewise, while presence of RT possums was associated with increased odds of a property being confirmed, BT possums were associated with decreased odds of a property being IS2404 detected. It is also important to note that all viable properties had RT possum feces present and thus adjustment for this factor was not included in the model. Increased property size and more alkaline soil were associated with being a confirmed property. Lower altitude was associated with a property being confirmed, while presence of power lines was associated with a property being IS2404 detected, confirmed and a case property.

### Return/follow-up property field surveys

A total of 27 properties were visited twice. The number of days between visits varied between 92 days (∼3 months) and 261 days (∼8.7 months), showing a right-skewed distribution with a median of 147 days (∼4.9 months; IQR = 92 days). Property status remained the same between visits for 19 (70.4%) properties based on IS2404 results, 22 (81.5%) for confirmed results and 23 (85.5%) for viability results (Table 5). At properties that remained positive, RT possum feces were the main sample type that remained positive at 10/14 (71%) properties for IS2404, 8/9 (89%) for confirmed and 2/2 (100%) for viable.

**Table 5:**
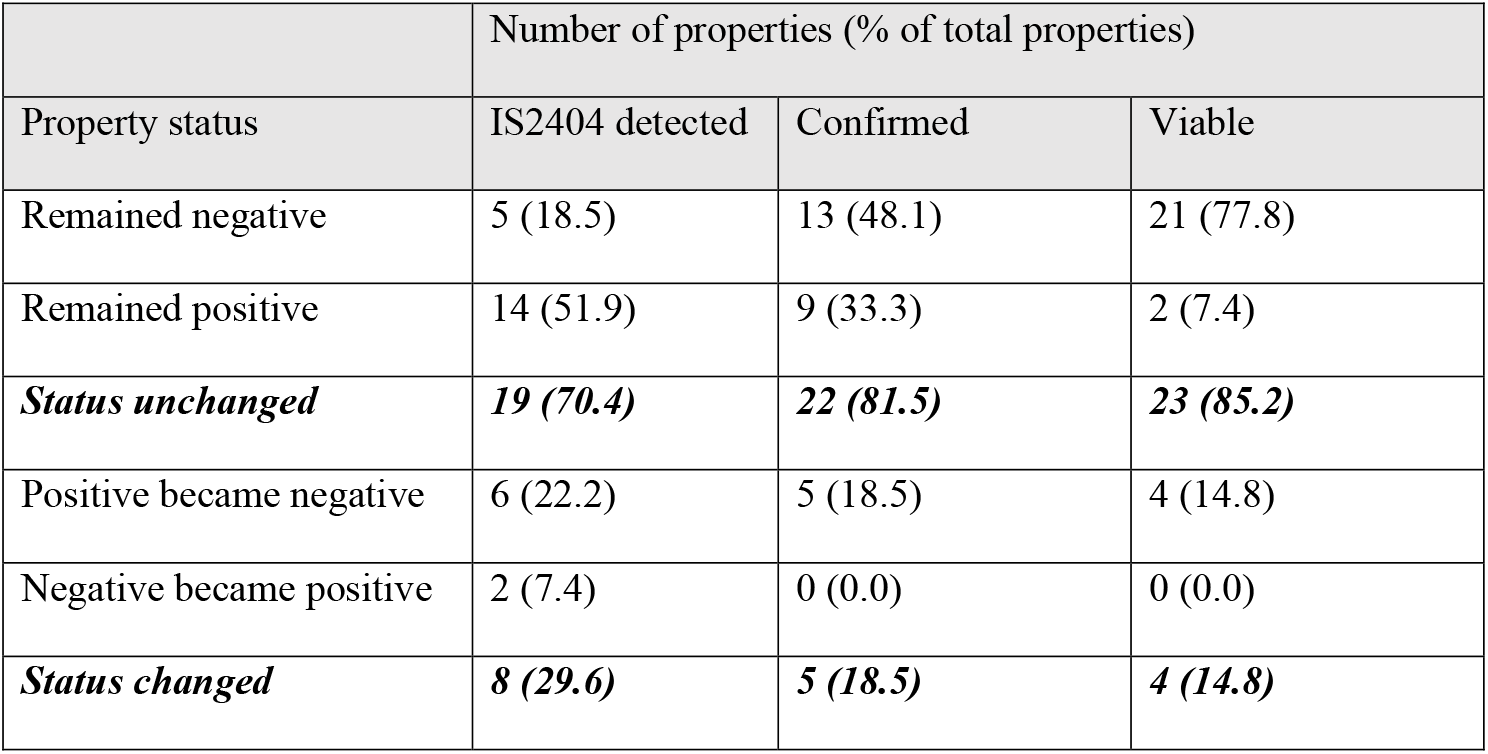
Property status between first and second field surveys by assay type

|  | Number of properties (% of total properties) |  |  |
| --- | --- | --- | --- |
| Property status | IS2404 detected | Confirmed | Viable |
| Remained negative | 5 (18.5) | 13 (48.1) | 21 (77.8) |
| Remained positive | 14 (51.9) | 9 (33.3) | 2 (7.4) |
| ***Status unchanged*** | ***19 (70.4)*** | ***22 (81.5)*** | ***23 (85.2)*** |
| Positive became negative | 6 (22.2) | 5 (18.5) | 4 (14.8) |
| Negative became positive | 2 (7.4) | 0 (0.0) | 0 (0.0) |
| ***Status changed*** | ***8 (29.6)*** | ***5 (18.5)*** | ***4 (14.8)*** |

Under the assumption that properties with positive status at both visits were positive for the entire period between the two sampling visits, we documented one property that remained IS2404 detected for at least 8.7 months, which was also the longest time between visits for any property. Of properties that remained IS2404 detected, half (7/14) had an interval between sampling of over six months. For nine confirmed properties, five properties remained confirmed for over six months, with the longest sampling interval of 7.9 months. The two properties that remained positive for the viability assay were positive for over six months. Only for the IS2404 assay did any property become positive, with two properties that were initially IS2404 negative becoming positive at the second visit (7.4%; Table 5); all other changes of property status were from positive at the first visit, to negative at the second visit (Table 5).

## Discussion

The findings from this study support the hypothesis that BU may be a zoonotic disease in Australia, with native mammals, specifically species of possum, acting as reservoir hosts (Carson et al., 2014; O’Brien et al., 2014). Consistent with previous findings, fecal samples were the sample type most commonly positive for MU and had the highest bacterial loads (Fyfe et al. 2010). This was also the sample type most likely to remain positive at return properties and the only sample type that appeared to contain viable bacteria. In Australia, numerous species of both native and introduced mammals including feces from RT possum, BT possum and rodents have tested positive for MU in the past, (Fyfe et al., 2010; Roltgen et al., 2017)) as also found in this study. In addition, in our study fecal samples from wild fox (*Vulpes vulpes*) and rabbit (*Oryctolagus cuniculus*) tested positive; a first report for both species, although laboratory rabbits have been infected experimentally (Norden & Linell, 1951). Fox feces collected during this study had the highest proportion of positives in IS2404 and confirmatory assays. However, RT possum feces were the sample type with the second highest proportion of positives by both these assays, the primary sample type positive by the viability assay and the sample type most commonly collected in this study (>28% of all samples collected). Only feces from RT possums, BT possums and a single fox were found to be viable, suggesting that all three species may be involved in the transmission of MU.

RT possums were more likely to be found at confirmed and viable properties and their feces were more likely to be positive for MU at case properties. Previous studies have also found a correlation between the geographic location of cases and the presence of positive possum feces (Carson et al., 2014; Fyfe et al 2010), supporting the reservoir host hypothesis. The findings from this study suggest that the presence of RT possums *per se* at a property does not increase the risk of the residents contracting BU, but the presence of RT possums positive for MU does. This intrinsically makes sense but requires effective communication to local residents to discourage the indiscriminate removal or translocation of possums, which are a protected species in Victoria. In the UK, removal of badgers as part of bovine tuberculosis (TB) control measures led to increased bovine TB prevalence in some regions. This was hypothesized to be because culling disrupted badger social organization, leading to long-distance movement and dispersal of individual badgers, resulting in increased TB transmission among badgers (Donnelly et al., 2003; Donnelly et al., 2006; Woodroffe et al., 2005). Increases in *Leptospira* carriage in rat populations subjected to indiscriminate lethal control methods in Vancouver, Canada have also been attributed to altered social structure and subsequent increases in aggressive interactions (Lee et al., 2018). As possums are territorial, removal or disturbance of individual resident animals impacts both social interactions and movement patterns (Matthews et al., 2004), which may in part help explain the shifting dynamics of this disease and the expansion of the Victorian endemic area.

Movement of MU into previously unaffected areas may also be facilitated by infected foxes, which demonstrate considerably larger home ranges than RT possums: individual foxes in a similar coastal habitat in New South Wales were found to have a mean home range of 135 ha (Meek & Saunders, 2000), compared to <1ha for RT possums (Lindenmayer et al., 2008). However, further research into the role of foxes in BU transmission is needed.

It is thought that MU can also persist outside of a vertebrate host, although the duration and the environmental conditions required have not been well defined (Bratschi et al., 2014; Garchitorena et al., 2015). Overall, soil was the second most commonly positive sample type, and particular soil characteristics were associated with positive properties. It is possible that the higher conductivity, salinity and alkalinity detected at these properties may enhance environmental survival of MU and/or aid transmission between hosts. The link between MU and slightly alkaline soil was unexpected as these bacteria have been associated with mildly acidic pH conditions in two aquatic communities in Cameroon (Garchitorena et al., 2015). Under laboratory conditions MU also appears to grow better at mildly acidic pH, although growth can also occur under mildly alkaline conditions (Portaels & Pattyn, 1982). It is possible that in soil (in contrast to water), other biotic factors may interact with pH to make alkaline conditions more favorable to MU. However, as only a minority of IS2404 positive soil samples were confirmed positive and none were considered viable, the detection of MU in soil may represent the presence of DNA from non-viable, degrading MU. If this is the case, then these environmental conditions may favor the preservation of MU DNA rather than bacterial survival. There seemed to be little association between MU and water at the scale analyzed in this study. Very few water sources returned IS2404 positive results and only a single water source was confirmed positive (from a bucket). This is consistent with previous environmental surveys in the region that also found low rates of MU positivity in soil and water (Fyfe et al 2010). There was also no association between property status and the number of water sources present or the presence of bore water. This suggests that in Victoria, water plays a limited role in determining the fine-scale distribution of MU.

Infection through puncturing injuries received from plants, biting insects and other objects contaminated with MU has also been hypothesized as a transmission pathway to humans (Street et al., 1991; Wallace et al., 2017). Relatively few insects were screened in this study making it difficult to assess associations with cases. However, one confirmed mosquito (*Aedes notoscriptus*)was detected in a case property, consistent with MU mosquito positivity reported in previous field surveys in this region (Johnson et al., 2007; Lavender et al., 2011). However, no association with mosquito or March fly presence and property status was observed. In addition, few plants tested positive by IS2404 for MU and only four samples were confirmed positive: one bromeliad (*Aechmea sp.*), one rose (*Rosa sp.*) and two yuccas (*Yucca sp.*). There was also no association between any of the most common spiky plant types and either case or positive properties. This suggests that plants, similar to water, are unlikely to be a common source of infection in Victoria. However, the presence of certain native plant species was associated with the presence of MU at properties. Coastal tea tree (*L. laevigatum*) and Moonah (*M. lanceolata*) are both indigenous to parts of the Mornington and Bellarine Peninsulas (Yugovic, 2002), and are utilized heavily by possum species for denning and as food sources (K. Blasdell, personal observation; Pahl, 1987). Interestingly, Moonahs were less likely to be found at case properties, although this may be due to the tendency of local residents (particularly those personally affected by MU) to discourage possums from visiting their properties through environmental modification due to the perception that possums are carriers of these bacteria. Certainly, the gardens of some properties visited by the researchers had been re-landscaped or modified by their owners post BU diagnosis (K Blasdell, personal observation). As gardens containing native or mixed vegetation were more likely to be positive for MU than those containing mainly non-native vegetation, this suggests that native environments may promote better survival of the bacteria, potentially because they appear to support denser populations of native mammalian hosts, such as possums.

To persist, possums require a suitable area of habitat containing sufficient resources. Habitat patches below a certain size (such as most urban and suburban gardens) are unlikely to provide these requirements unless they are well connected to other similar patches (Goddard et al., 2010). For example, individual BT possums in Melbourne, Australia regularly foraged in several residential gardens despite denning in urban forest fragments (Harper et al., 2005), whilst in New Zealand, BT possum occupancy of urban gardens decreased with increasing housing density and decreasing green cover (Adams et al., 2013). Assuming that RT possums respond in a similar way, this may explain the association between larger properties and positive status. However, this could also be a geographic effect, as properties surveyed in the Mornington Peninsula (the current epicenter of BU in Victoria/Australia) were larger than those surveyed closer to Melbourne (Bayside area). Powerlines were more likely to be found at IS2404 detected, confirmed and case properties. As possums regularly use power lines to travel around urban areas (K Blasdell, personal observation), this feature might promote connectivity between properties and facilitate the presence of these potential hosts. Similar to our study, BU prevalence was found to increase with decreasing elevation in Benin, with the authors proposing that MU survival might be promoted by the wetter conditions often found at lower altitudes (Sopoh et al., 2011).

Although return surveys where only conducted at a small proportion of properties, the findings suggest that MU bacteria can remain at a specific location for a considerable period of time (>6 months). This has also been found in Cameroon, where a village water source remained positive for over two years (Bratschi et al., 2014). However, as each property was only sampled at two time points, it is possible that undetected changes may have occurred at properties during that interval, and additionally that a property might remain positive for MU for longer than the maximum 8.7 months observed here. Although it is unknown what factors changed between sampling points for those properties where MU status did alter, some environmental changes were observed at some of these properties that may have impacted the presence and survivability of MU. For example, at one property that became IS2404 positive at the return visit, de-vegetation and construction of a new house on the neighboring plot, which had previously been vacant and covered in native flora, may have resulted in the movement of infected wildlife onto the sampled plot. Most properties changed from positive to negative, which may suggest that the environmental disease risk in this region decreased slightly over the study period. At individual properties this may be because the resident infected possum (or other host) dies and is replaced by a non-infected individual, although this requires further exploration. However, two properties did become positive by IS2404, demonstrating this is a dynamic situation.

This study’s findings cast some light on (1) what the ‘ideal property’ for MU presence looks like and (2) how this differs from what the ‘ideal’ case property looks like. The results suggest that the average MU property is a larger property located at lower altitudes with soil that is slightly alkaline. It has overhead powerlines and contains native vegetation, particularly coastal tea trees, which in turn support a healthy population of RT possums. In contrast, the ‘ideal’ case property has overhead powerlines present, is less likely to contain Moonahs, but more likely to contain MU-positive wild mammals, especially RT possums. Further longitudinal investigation is required to understand these differences in environmental associations between properties where MU was detected and properties where human cases occurred. It should be noted that although there was a clear association between the presence of infected RT possums and human cases, MU-positive RT possum feces and other samples were also found at many control properties. Human behavioral impacts on BU disease risk will be assessed through the analysis of the questionnaires collected as part of this case-control study (results to be published separately).

Our findings provide additional evidence to support the hypothesis that MU is a zoonotic pathogen, at least in the Victorian endemic area. Although RT possums are clearly the strongest reservoir host candidate, the high proportion of fox feces that were IS2404 and confirmed positive, along with evidence of viability in one sample, indicate the foxes may also contribute to the circulation of this pathogen. Further research is needed to explore whether the more mobile fox could introduce MU to new areas.

## Supporting information

STROBE checklist

COI disclosure

## Data Availability

All data produced in the present study are available upon reasonable request to the authors

## Acknowledgments

We gratefully acknowledge Loretta Vaughen for help with accessing the VPHS database. We also thank all who assisted with field collections and questionnaire deployment, and all of the project participants, with particular thanks to those who allowed us to conduct field surveys at their properties.

## Author contributions

KRB created the study design, collected, analyzed and interpreted the data and drafted the manuscript. BM analyzed and interpreted the data and drafted the manuscript. DPO’B assisted with the study design, interpreted the data and drafted the manuscript. MT, SC and JG assisted with the study design and collected the data. VB, MD and PTM assisted with the study design, collected, analyzed and interpreted the data. IS, KBG, ET and SEL assisted with the study design, interpreted the data and drafted the manuscript. ECH and NW interpreted the data and drafted the manuscript. TPS and EA assisted with the study design, interpreted the data and drafted the manuscript. All authors reviewed and edited the manuscript.

## Supplemental figure legends

Supplemental Figure 1: Key indigenous (panels A-D) and non-indigenous (panel E) plants recorded for each property. A – *Melaleuca lanceolata* (Moonah/black paperbark); B - *Leptospernum laevigatum* (coastal tea tree); C - *Leucopogon parviflorus* (coast beard heath/native currant); D – *Allocasuarina verticillata/littoralis* (Drooping and black sheoaks); E – *Pittosporum* spp. (cheesewoods).

Supplemental Figure 2: Flow diagram for environmental property surveys and sample collection.

Supplemental Figure 3: Example of a property outline with locations of key features and sample locations marked. Yellow sticky traps (YST) were placed at the majority of properties for additional insect capture. Results from these traps will be reported in a separate publication.

Supplemental Figure 4: Examples of plant samples collected; Panel A – selection of ‘spiky’ plants sampled; Panel B – selection of fruits with evidence of mammalian gnaw marks.

Supplemental Figure 5: Directed acyclic graph (DAG) used for assessing the potential for confounding by covariates and for identifying the appropriate confounders to be included in each adjusted model.

Supplemental Figure 6: Map of affected area, illustrating the MU status by suburb. Suburbs containing at least one ‘viable’ property were classified as viable. Suburbs without ‘viable’ properties but with at least one ‘confirmed’ property were classified as confirmed. Suburbs without ‘viable’ or ‘confirmed’ properties but with at least one ‘IS2404 detected’ property were classified as ‘IS2404 detected’. Suburbs without any ‘IS2404 detected’ properties were classified as negative. N.B. Geographical boundaries are not available by postcode and some postcodes contain more than one suburb.

