## Supplemental Table 1 for "Environmental risk factors associated with the presence of *Mycobacterium ulcerans* in Victoria, Australia"

Supplemental Table 1 – Univariate statistics: Environmental characteristics demonstrating significant relationships with at least one property type with numbers (and percentages) or average values shown by property type. Significant relationships are shown in bold.

| **Environmental characteristic** | **Test used** | **Property type** | | | | | | | | |
| --- | --- | --- | --- | --- | --- | --- | --- | --- | --- | --- |
|  |  | *IS2404* | | *Confirmed* | | *Viable* | | | *Case* | |
|  |  | Positive: n = 157 (68.3%) | Negative: n = 73 (31.7%) | Confirmed: n = 103 (44.8%) | Unconfirmed: n =127 (55.2%) | Viable: n = 46 (20.0%) | | Non-viable: n = 184 (80.0%) | Case: n = 115 (50.0%) | Control: n = 115 (50.0%) |
| **Garden type:**  NN (n = 75)  M (n = 82)  N (n = 73) | Chi-square | 37 (23.6%)  62 (39.5%)  58 (36.9%) | 38 (52.1%)  20 (27.4%)  15 (20.5%) | 21 (20.4%)  38 (36.9%)  44 (42.7%) | 54 (42.5%)  44 (34.6%)  29 (22.8%) | 8 (17.4%)  17 (37.0%)  21 (45.7%) | | 67 (36.4%)  65 (35.3%)  52 (28.3%) | 35 (30.4%)  45 (39.1%)  35 (30.4%) | 40 (34.8%)  37 (32.2%)  38 (33.0%) |
|  |  | **P<0.001** | | **P<0.001** | | **P<0.05** | | | N/S | |
| **Presence of *Melaleuca lanceolata*** | Chi-square | 87 (55.4%) | 24 (32.9%) | 65 (63.1%) | 46 (36.2%) | 33 (71.7%) | | 78 (42.4%) | 47 (40.9%) | 64 (55.7%) |
|  |  | **P<0.01** | | **P<0.001** | | **P<0.001** | | | **P<0.05** | |
| **Presence of *Leptospernum laevigatum*** | Chi-square | 106 (67.5%) | 29 (39.7%) | 79 (76.7%) | 56 (44.1%) | 37 (80.4%) | | 98 (53.3%) | 70 (60.9%) | 65 (56.5%) |
|  |  | **P<0.001** | | **P<0.001** | | **P<0.001** | | | N/S | |
| **Presence of *Leucopogon parviflorus*** | Chi-square | 60 (38.2%) | 19 (26.0%) | 40 (38.8%) | 39 (30.7%) | 22 (47.8%) | | 57 (31.0%) | 33 (28.7%) | 46 (40.0%) |
|  |  | N/S (P<0.1) | | N/S | | **P<0.05** | | | N/S (P<0.1) | |
| **Presence of *Allocasuarina verticallata/ littoralis*** | Chi-square | 41 (26.1%) | 11 (15.1%) | 18 (17.5%) | 23 (18.1%) | 7 (15.2%) | 34 (18.55) | | 17 (14.8%) | 24 (20.9%) |
|  |  | N/S (P<0.1) | | N/S | | N/S | | | N/S | |
| **Presence of *Pittosporum* spp.** | Chi-square | 43 (27.4%) | 32 (43.8%) | 32 (31.1%) | 43 (33.9%) | 13 (28.3%) | | 62 (33.7%) | 32 (27.8%) | 43 (37.4%) |
|  |  | **P<0.05** | | N/S | | N/S | | | N/S | |
| **Presence of spiky roses** | Chi-square | 75 (47.8%) | 39 (53.4%) | 45 (43.7%) | 69 (54.3%) | 18 (39.1%) | | 96 (52.2%) | 55 (47.8%) | 59 (51.3%) |
|  |  | N/S | | N/S | | N/S | | | N/S | |
| **Presence of spiky citrus** | Chi-square | 62 (39.5%) | 29 (39.7%) | 41 (39.8%) | 50 (39.4%) | 18 (39.1%) | | 73 (39.7%) | 49 (42.6%) | 42 (36.5%) |
|  |  | N/S | | N/S | | N/S | | | N/S | |
| **Presence of spiky succulents** | Chi-square | 47 (29.9%) | 13 (17.8%) | 31 (30.1%) | 29 (22.8%) | 9 (19.6%) | | 51 (27.7%) | 33 (28.7%) | 27 (23.5%) |
|  |  | N/S (P<0.1) | | N/S | | N/S | | | N/S | |
| **Presence of spiky yuccas** | Chi-square | 45 (28.7%) | 26 (35.6%) | 32 (31.1%) | 39 (30.7%) | 16 (34.8%) | | 55 (29.9%) | 32 (27.0%) | 39 (33.9%) |
|  |  | N/S | | N/S | | N/S | | | N/S | |
| **Presence of spiky bromeliads** | Chi-square | 20 (12.7%) | 10 (13.7%) | 15 (14.6%) | 15 (11.8%) | 7 (15.2%) | | 23 (12.5%) | 19 (16.5%) | 11 (9.6%) |
|  |  | N/S | | N/S | | N/S | | | N/S | |
| **Presence of spiky cacti** | Chi-square | 22 (14.0%) | 12 (16.4%) | 16 (15.5%) | 18 (14.2%) | 5 (10.9%) | | 29 (15.8%) | 16 (13.9%) | 18 (15.7%) |
|  |  | N/S | | N/S | | N/S | | | N/S | |
| **Presence of spiky cycads** | Chi-square/  ^+^Fishers Exact | 15 (9.6%) | 7 (9.6%) | 8 (7.8%) | 14 (11.0%) | 3 (6.5%) | | 19 (10.3%) | 10 (8.7%) | 12 (10.4%) |
|  |  | N/S | | N/S | | ^+^N/S | | | N/S | |
| **Presence of RT possum faeces** | Chi-square | 150 (95.5%) | 62 (84.9%) | 102 (99.0%) | 110 (86.6%) | 46 (100.0%) | | 166 (90.2%) | 106 (92.2%) | 106 (92.2%) |
|  |  | **P<0.01** | | **P<0.001** | | **P<0.05** | | | N/S | |
| **Presence of BT possum faeces** | Chi-square | 44 (28.0%) | 34 (46.6%) | 30 (29.1%) | 48 (37.8%) | 13 (28.3%) | | 65 (35.3%) | 39 (33.9%) | 39 (33.9%) |
|  |  | **P<0.01** |  | N/S | | N/S | | | N/S | |
| **Presence of rodent faeces** | Chi-square | 56 (35.7%) | 33 (45.2%) | 32 (31.1%) | 57 (44.9%) | 13 (28.3%) | | 76 (41.3%) | 47 (40.9%) | 42 (36.5%) |
|  |  | N/S | | **P<0.05** | | N/S | | | N/S | |
| **Presence of fox faeces** | Chi-square/  ^+^Fishers Exact | 14 (8.9%) | 3 (4.1%) | 10 (9.7%) | 7 (5.5%) | 3 (6.5%) | | 14 (7.1%) | 8 (7.0%) | 8 (7.0%) |
|  |  | N/S | | N/S | | ^+^N/S | | | N/S | |
| **Presence of rabbit faeces** | Chi-square | 9 (5.7%) | 1 (1.4%) | 7 (6.8%) | 3 (2.4%) | 5 (10.9%) | | 5 (2.7%) | 2 (1.7%) | 8 (7.0%) |
|  |  | N/S | | N/S | | **P<0.05** | | | N/S | |
| **Presence of adult mosquitoes** | Chi-square | 50 (31.8%) | 25 (34.2%) | 30 (29.1%) | 45 (35.4%) | 10 (21.7%) | | 65 (35.3%) | 41 (35.7%) | 34 (29.6%) |
|  |  | N/S | | N/S | | N/S | | | N/S | |
| **Presence of March flies** | Fishers Exact/ ^#^Chi-square | 9 (5.7%) | 3 (4.1%) | 4 (3.9%) | 8 (6.3%) | 1 (2.2%) | | 11 (6.0%) | 6 (5.2%) | 6 (5.2%) |
|  |  | N/S | | N/S | | N/S | | | N/S^#^ | |
| **Presence of mosquito larvae** | Chi-square | 76 (48.4%) | 32 (43.8%) | 48 (46.6%) | 60 (47.2%) | 18 (39.1%) | | 90 (48.9%) | 58 (50.4%) | 50 (43.5%) |
|  |  | N/S | | N/S | | N/S | | | N/S | |
| **Presence of bore water** | Chi-square | 32 (20.4%) | 9 (12.3%) | 24 (23.3%) | 17 (13.4%) | 12 (26.1%) | | 29 (15.8%) | 23 (20.0%) | 18 (15.7%) |
|  |  | N/S | | N/S (P<0.1) | | N/S | | | N/S | |
| **Number of water sources** | T-test | 4 | 5 | 4 | 5 | 4 | | 5 | 4 | 5 |
|  |  | N/S | | N/S | | N/S | | | N/S | |
| **Property size (m^2^)** | T-test | 1042 | 800 | 1141 | 823 | 1202 | | 906 | 893 | 1037 |
|  |  | **P<0.05** | | **P<0.01** | | **P<0.05** | | | N/S | |
| **Presence of powerlines** | Chi-square | 131 (83.4%) | 52 (71.2%) | 88 (85.4%) | 95 (74.8%) | 39 (84.8%) | | 144 (78.3%) | 101 (87.8%) | 82 (71.3%) |
|  |  | **P<0.05** | | **P = 0.05** | | N/S | | | **P<0.01** | |
| **Powerlines attached to the house** | Chi-square | 100 (63.7%) | 47 (64.4%) | 70 (68.0%) | 77 (60.6%) | 33 (71.7%) | | 114 (62.0%) | 81 (70.4%) | 66 (57.4%) |
|  |  | N/S | | N/S | | N/S | | | **P<0.05** | |
| **Powerlines along edge of property** | Chi-square | 74 (47.1%) | 20 (27.4%) | 47 (45.6%) | 47 (37.0%) | 18 (39.1%) | | 76 (41.3%) | 58 (50.4%) | 36 (31.3%) |
|  |  | **P<0.01** | | N/S | | N/S | | | **P<0.01** | |
| **Altitude (m)** | T-test | 17.5 | 23.8 | 15.6 | 22.6 | 16.4 | | 20.3 | 19.0 | 19.9 |
|  |  | **P<0.05** | | **P<0.01** | | N/S | | | N/S | |
| **Soil bulk density (g/cm^3^)** | T-test | 0.94 | 0.94 | 0.94 | 0.94 | 0.92 | | 0.95 | 0.96 | 0.93 |
|  |  | N/S | | N/S | | N/S | | | N/S | |
| **Soil conductivity (µS/cm)** | T-test | 149.9 | 115.8 | 160.9 | 121.4 | 182.4 | | 128.3 | 146.7 | 131.4 |
|  |  | **P<0.05** | | **P<0.01** | | **P<0.01** | | | N/S | |
| **Soil salinity class*:**  NS (n = 4)  SlS (n = 21)  MS (n = 86)  HS (n = 173)  SeS (n = 107)  ES (n = 69) | Chi-square | 2 (0.6%)  6 (1.9%)  54 (17.2%)  123 (39.2%)  77 (24.5%)  52 (16.6%) | 2 (1.4%)  15 (10.3%)  32 (21.9%)  50 (34.2%)  30 (20.5%)  17 (11.6%) | 1 (0.5%)  2 (1.0%)  28 (13.6%)  84 (40.8%)  53 (25.7%)  38 (18.4%) | 3 (1.2%)  19 (7.5%)  58 (22.8%)  89 (35.0%)  54 (21.3%)  31 (12.2%) | 1 (1.1%)  1 (1.1%)  12 (13.0%)  38 (41.3%)  19 (20.7%)  21 (22.8%) | | 3 (0.8%)  20 (5.4%)  74 (20.1%)  135 (36.7%)  88 (23.9%)  48 (13.0%) | 1 (0.4%)  8 (3.5%)  40 (17.4%)  88 (38.3%)  56 (24.3%)  37 (16.1%) | 3 (1.3%)  13 (5.7%)  46 (20.0%)  85 (40.0%)  51 (22.2%)  32 (13.9%) |
|  |  | **P<0.01** | | **P<0.001** | | N/S (P<0.1) | | | N/S | |
| **Soil pH** | T-test | pH 8.0 | pH 7.5 | pH 8.1 | pH 7.6 | pH 8.2 | | pH 7.7 | pH 7.8 | pH 7.8 |
|  |  | **P<0.001** | | **P<0.001** | | **P<0.001** | | | N/S | |
| **Soil texture type***:  S (n = 25)  SL (n = 376)  L (n = 40)  C (n = 19) | Chi-square | 12 (3.8%)  266 (84.7%)  26 (8.2%)  10 (3.2%) | 13 (8.9%)  115 (75.3%)  14 (9.6%)  9 (6.1%) | 9 (4.4%)  180 (87.4%)  14 (6.8%)  3 (1.5%) | 16 (6.3%)  196 (77.2%)  26 (10.2%)  16 (6.3%) | 3 (3.3%)  81 (88.0%)  6 (6.3%)  2 (2.1%) | | 22 (6.0%)  295 (80.2%)  34 (9.2%)  17 (4.6%) | 11 (4.8%)  192 (83.5%)  18 (7.8%)  9 (3.9%) | 14 (6.1%)  184 (80.0%)  22 (9.6%)  10 (4.4%) |
|  |  | **P<0.05** | | **P<0.05** | | N/S | | | N/S | |

*Key: NN = Non-native; M = Mixed; N = Native; NS = Non saline; SlS = Slightly saline; MS = Moderately saline; HS = Highly saline; SeS = Severely saline; ES = Extremely saline; S = Sand; SL = Sandy Loam; L = Loam; C = Clay/clay loam. N.B. Two soil samples were collected at each property, therefore numbers are twice the value of all other categories.
