## Supplemental Material for "Environmental risk factors associated with the presence of *Mycobacterium ulcerans* in Victoria, Australia"

**Field Collection Sheet**

**Residence ID:**

**Field staff: _____________________________________________________________**

**Date:	______________	GPS coordinates: ___________________**

**Altitude (m): ________	Approx size (m^2^): ___________	Approx age:	______**

**Garden type:	_____________________	Housing density: ____________________**

**Powerlines:** None 🞏	Attached 🞏	Along edge 🞏	Other 🞏

**Insect collection methods used:** Aspirator	🞏	Sticky traps 🞏	Other	🞏

_______________________________________________________________________

**Vegetation**

| **Latin name** | **Common name(s)** | **Present** |
| --- | --- | --- |
| *Melaleuca lanceolate* | Moonah / black paperbark |  |
| *Leptospernum laevigatum* | Coastal tea tree |  |
| *Leucopogon parviflorus* | Coast beard-heath / native currant |  |
| *Allocasuarina verticillata/littoralis* | Drooping sheoak |  |
| *Pittosporum* spp. | Pittosporums (larger varieties) |  |

**Photos/videos:** 180°/360° photos 🞏	Video 🞏	Other 🞏 _______________

**Location where participant spends most time (for larger properties): ____________**

**_______________________________________________________________________**

**Plants native animals are eating: ___________________________________________**

**Other info: _____________________________________________________________**

**________________________________________________________________________**

**________________________________________________________________________**

**________________________________________________________________________**

**________________________________________________________________________**

**Sample details**

***Sample 1***	Location ___________________________________	Photo ID ___________

**Soil**	🞏	Colour	___________	Temp	________	pH _______

Conductivity (uS/cm) _______	EC/Salinity ________	Texture ____________

**Water**	🞏	Source type _______________________________	WS no. ______

Mosquito larvae collected 🞏	Collection technique _____________________

**Insect**	🞏	Midge 🞏	Marsh fly 🞏	Mosquito 🞏	Other 🞏	Collection technique ____________________

**Plant**	🞏	Type _______________________________	Spiky 🞏	Food source 🞏

**Faeces**	🞏	Species _____________________________________

***Sample 2***	Location ___________________________________	Photo ID ___________

**Soil**	🞏	Colour	___________	Temp	________	pH _______

Conductivity (uS/cm) _______	EC/Salinity ________	Texture ____________

**Water**	🞏	Source type _______________________________	WS no. ______

Mosquito larvae collected 🞏	Collection technique _____________________

**Insect**	🞏	Midge 🞏	Marsh fly 🞏	Mosquito 🞏	Other 🞏	Collection technique ____________________

**Plant**	🞏	Type _______________________________	Spiky 🞏	Food source 🞏

**Faeces**	🞏	Species _____________________________________

***Sample 3***	Location ___________________________________	Photo ID ___________

**Soil**	🞏	Colour	___________	Temp	________	pH _______

Conductivity (uS/cm) _______	EC/Salinity ________	Texture ____________

**Water**	🞏	Source type _______________________________	WS no. ______

Mosquito larvae collected 🞏	Collection technique _____________________

**Insect**	🞏	Midge 🞏	Marsh fly 🞏	Mosquito 🞏	Other 🞏	Collection technique ____________________

**Plant**	🞏	Type _______________________________	Spiky 🞏	Food source 🞏

**Faeces**	🞏	Species _____________________________________

***Sample 4***	Location ___________________________________	Photo ID ___________

**Soil**	🞏	Colour	___________	Temp	________	pH _______

Conductivity (uS/cm) _______	EC/Salinity ________	Texture ____________

**Water**	🞏	Source type _______________________________	WS no. ______

Mosquito larvae collected 🞏	Collection technique _____________________

**Insect**	🞏	Midge 🞏	Marsh fly 🞏	Mosquito 🞏	Other 🞏	Collection technique ____________________

**Plant**	🞏	Type _______________________________	Spiky 🞏	Food source 🞏

**Faeces**	🞏	Species _____________________________________

***Sample 5***	Location ___________________________________	Photo ID ___________

**Soil**	🞏	Colour	___________	Temp	________	pH _______

Conductivity (uS/cm) _______	EC/Salinity ________	Texture ____________

**Water**	🞏	Source type _______________________________	WS no. ______

Mosquito larvae collected 🞏	Collection technique _____________________

**Insect**	🞏	Midge 🞏	Marsh fly 🞏	Mosquito 🞏	Other 🞏	Collection technique ____________________

**Plant**	🞏	Type _______________________________	Spiky 🞏	Food source 🞏

**Faeces**	🞏	Species _____________________________________

***Sample 6***	Location ___________________________________	Photo ID ___________

**Soil**	🞏	Colour	___________	Temp	________	pH _______

Conductivity (uS/cm) _______	EC/Salinity ________	Texture ____________

**Water**	🞏	Source type _______________________________	WS no. ______

Mosquito larvae collected 🞏	Collection technique _____________________

**Insect**	🞏	Midge 🞏	Marsh fly 🞏	Mosquito 🞏	Other 🞏	Collection technique ____________________

**Plant**	🞏	Type _______________________________	Spiky 🞏	Food source 🞏

**Faeces**	🞏	Species _____________________________________

***Sample 7***	Location ___________________________________	Photo ID ___________

**Soil**	🞏	Colour	___________	Temp	________	pH _______

Conductivity (uS/cm) _______	EC/Salinity ________	Texture ____________

**Water**	🞏	Source type _______________________________	WS no. ______

Mosquito larvae collected 🞏	Collection technique _____________________

**Insect**	🞏	Midge 🞏	Marsh fly 🞏	Mosquito 🞏	Other 🞏	Collection technique ____________________

**Plant**	🞏	Type _______________________________	Spiky 🞏	Food source 🞏

**Faeces**	🞏	Species _____________________________________

***Sample 8***	Location ___________________________________	Photo ID ___________

**Soil**	🞏	Colour	___________	Temp	________	pH _______

Conductivity (uS/cm) _______	EC/Salinity ________	Texture ____________

**Water**	🞏	Source type _______________________________	WS no. ______

Mosquito larvae collected 🞏	Collection technique _____________________

**Insect**	🞏	Midge 🞏	Marsh fly 🞏	Mosquito 🞏	Other 🞏	Collection technique ____________________

**Plant**	🞏	Type _______________________________	Spiky 🞏	Food source 🞏

**Faeces**	🞏	Species _____________________________________

***Sample 9***	Location ___________________________________	Photo ID ___________

**Soil**	🞏	Colour	___________	Temp	________	pH _______

Conductivity (uS/cm) _______	EC/Salinity ________	Texture ____________

**Water**	🞏	Source type _______________________________	WS no. ______

Mosquito larvae collected 🞏	Collection technique _____________________

**Insect**	🞏	Midge 🞏	Marsh fly 🞏	Mosquito 🞏	Other 🞏	Collection technique ____________________

**Plant**	🞏	Type _______________________________	Spiky 🞏	Food source 🞏

**Faeces**	🞏	Species _____________________________________

***Sample 10***	Location ___________________________________	Photo ID ___________

**Soil**	🞏	Colour	___________	Temp	________	pH _______

Conductivity (uS/cm) _______	EC/Salinity ________	Texture ____________

**Water**	🞏	Source type _______________________________	WS no. ______

Mosquito larvae collected 🞏	Collection technique _____________________

**Insect**	🞏	Midge 🞏	Marsh fly 🞏	Mosquito 🞏	Other 🞏	Collection technique ____________________

**Plant**	🞏	Type _______________________________	Spiky 🞏	Food source 🞏

**Faeces**	🞏	Species _____________________________________

***Sample 11***	Location ___________________________________	Photo ID ___________

**Soil**	🞏	Colour	___________	Temp	________	pH _______

Conductivity (uS/cm) _______	EC/Salinity ________	Texture ____________

**Water**	🞏	Source type _______________________________	WS no. ______

Mosquito larvae collected 🞏	Collection technique _____________________

**Insect**	🞏	Midge 🞏	Marsh fly 🞏	Mosquito 🞏	Other 🞏	Collection technique ____________________

**Plant**	🞏	Type _______________________________	Spiky 🞏	Food source 🞏

**Faeces**	🞏	Species _____________________________________

***Sample 12***	Location ___________________________________	Photo ID ___________

**Soil**	🞏	Colour	___________	Temp	________	pH _______

Conductivity (uS/cm) _______	EC/Salinity ________	Texture ____________

**Water**	🞏	Source type _______________________________	WS no. ______

Mosquito larvae collected 🞏	Collection technique _____________________

**Insect**	🞏	Midge 🞏	Marsh fly 🞏	Mosquito 🞏	Other 🞏	Collection technique ____________________

**Plant**	🞏	Type _______________________________	Spiky 🞏	Food source 🞏

**Faeces**	🞏	Species _____________________________________

***Sample 13***	Location ___________________________________	Photo ID ___________

**Soil**	🞏	Colour	___________	Temp	________	pH _______

Conductivity (uS/cm) _______	EC/Salinity ________	Texture ____________

**Water**	🞏	Source type _______________________________	WS no. ______

Mosquito larvae collected 🞏	Collection technique _____________________

**Insect**	🞏	Midge 🞏	Marsh fly 🞏	Mosquito 🞏	Other 🞏	Collection technique ____________________

**Plant**	🞏	Type _______________________________	Spiky 🞏	Food source 🞏

**Faeces**	🞏	Species _____________________________________

***Sample 14***	Location ___________________________________	Photo ID ___________

**Soil**	🞏	Colour	___________	Temp	________	pH _______

Conductivity (uS/cm) _______	EC/Salinity ________	Texture ____________

**Water**	🞏	Source type _______________________________	WS no. ______

Mosquito larvae collected 🞏	Collection technique _____________________

**Insect**	🞏	Midge 🞏	Marsh fly 🞏	Mosquito 🞏	Other 🞏	Collection technique ____________________

**Plant**	🞏	Type _______________________________	Spiky 🞏	Food source 🞏

**Faeces**	🞏	Species _____________________________________

***Sample 15***	Location ___________________________________	Photo ID ___________

**Soil**	🞏	Colour	___________	Temp	________	pH _______

Conductivity (uS/cm) _______	EC/Salinity ________	Texture ____________

**Water**	🞏	Source type _______________________________	WS no. ______

Mosquito larvae collected 🞏	Collection technique _____________________

**Insect**	🞏	Midge 🞏	Marsh fly 🞏	Mosquito 🞏	Other 🞏	Collection technique ____________________

**Plant**	🞏	Type _______________________________	Spiky 🞏	Food source 🞏

**Faeces**	🞏	Species _____________________________________

***Sample 16***	Location ___________________________________	Photo ID ___________

**Soil**	🞏	Colour	___________	Temp	________	pH _______

Conductivity (uS/cm) _______	EC/Salinity ________	Texture ____________

**Water**	🞏	Source type _______________________________	WS no. ______

Mosquito larvae collected 🞏	Collection technique _____________________

**Insect**	🞏	Midge 🞏	Marsh fly 🞏	Mosquito 🞏	Other 🞏	Collection technique ____________________

**Plant**	🞏	Type _______________________________	Spiky 🞏	Food source 🞏

**Faeces**	🞏	Species _____________________________________

***Sample 17***	Location ___________________________________	Photo ID ___________

**Soil**	🞏	Colour	___________	Temp	________	pH _______

Conductivity (uS/cm) _______	EC/Salinity ________	Texture ____________

**Water**	🞏	Source type _______________________________	WS no. ______

Mosquito larvae collected 🞏	Collection technique _____________________

**Insect**	🞏	Midge 🞏	Marsh fly 🞏	Mosquito 🞏	Other 🞏	Collection technique ____________________

**Plant**	🞏	Type _______________________________	Spiky 🞏	Food source 🞏

**Faeces**	🞏	Species _____________________________________

***Sample 18***	Location ___________________________________	Photo ID ___________

**Soil**	🞏	Colour	___________	Temp	________	pH _______

Conductivity (uS/cm) _______	EC/Salinity ________	Texture ____________

**Water**	🞏	Source type _______________________________	WS no. ______

Mosquito larvae collected 🞏	Collection technique _____________________

**Insect**	🞏	Midge 🞏	Marsh fly 🞏	Mosquito 🞏	Other 🞏	Collection technique ____________________

**Plant**	🞏	Type _______________________________	Spiky 🞏	Food source 🞏

**Faeces**	🞏	Species _____________________________________

***Sample 19***	Location ___________________________________	Photo ID ___________

**Soil**	🞏	Colour	___________	Temp	________	pH _______

Conductivity (uS/cm) _______	EC/Salinity ________	Texture ____________

**Water**	🞏	Source type _______________________________	WS no. ______

Mosquito larvae collected 🞏	Collection technique _____________________

**Insect**	🞏	Midge 🞏	Marsh fly 🞏	Mosquito 🞏	Other 🞏	Collection technique ____________________

**Plant**	🞏	Type _______________________________	Spiky 🞏	Food source 🞏

**Faeces**	🞏	Species _____________________________________

***Sample 20***	Location ___________________________________	Photo ID ___________

**Soil**	🞏	Colour	___________	Temp	________	pH _______

Conductivity (uS/cm) _______	EC/Salinity ________	Texture ____________

**Water**	🞏	Source type _______________________________	WS no. ______

Mosquito larvae collected 🞏	Collection technique _____________________

**Insect**	🞏	Midge 🞏	Marsh fly 🞏	Mosquito 🞏	Other 🞏	Collection technique ____________________

**Plant**	🞏	Type _______________________________	Spiky 🞏	Food source 🞏

**Faeces**	🞏	Species _____________________________________
