## Supplementary figures and images for "Environmental risk factors associated with the presence of *Mycobacterium ulcerans* in Victoria, Australia"

### Supplemental Figure 1

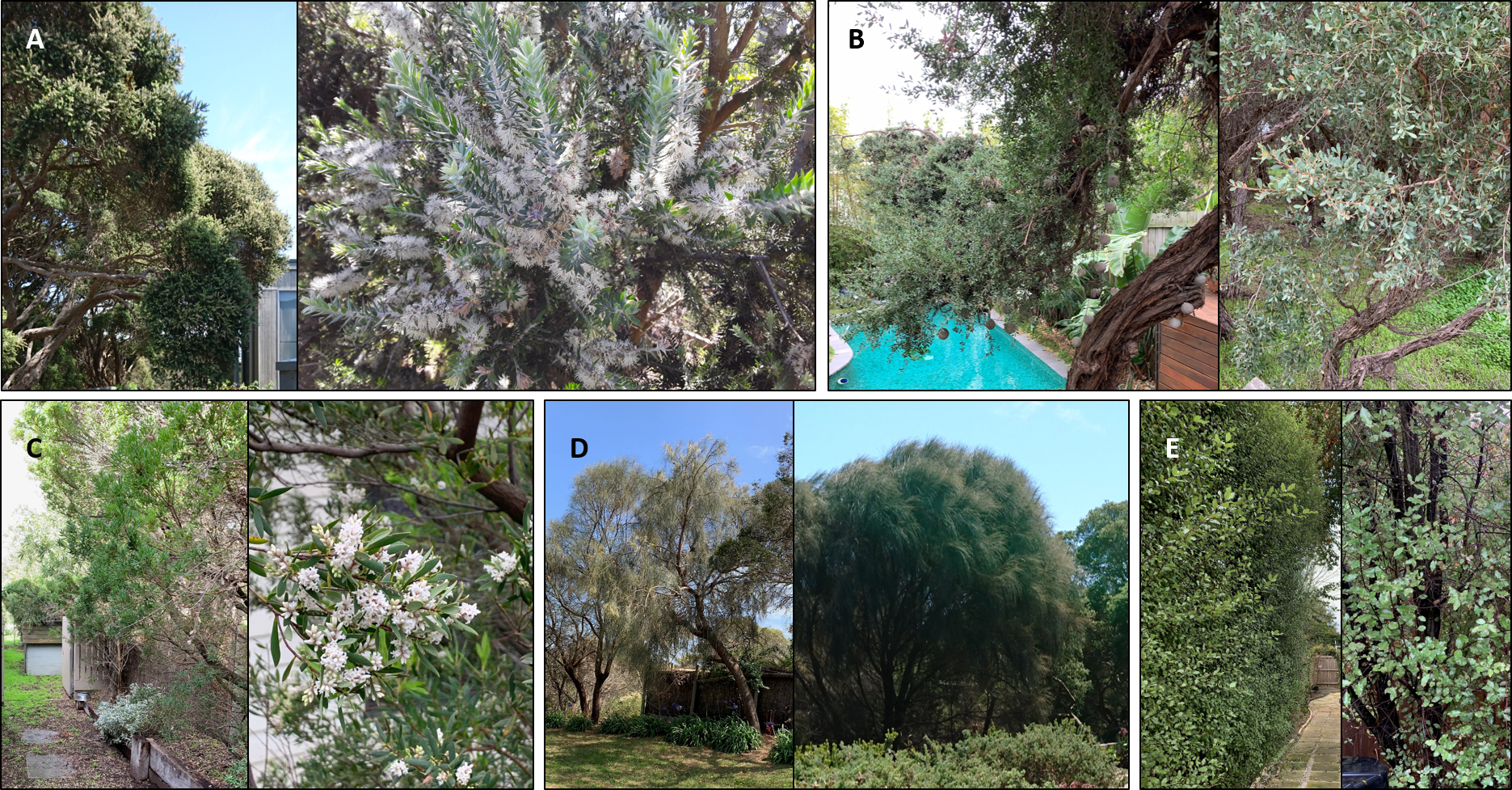

### Supplemental Figure 2

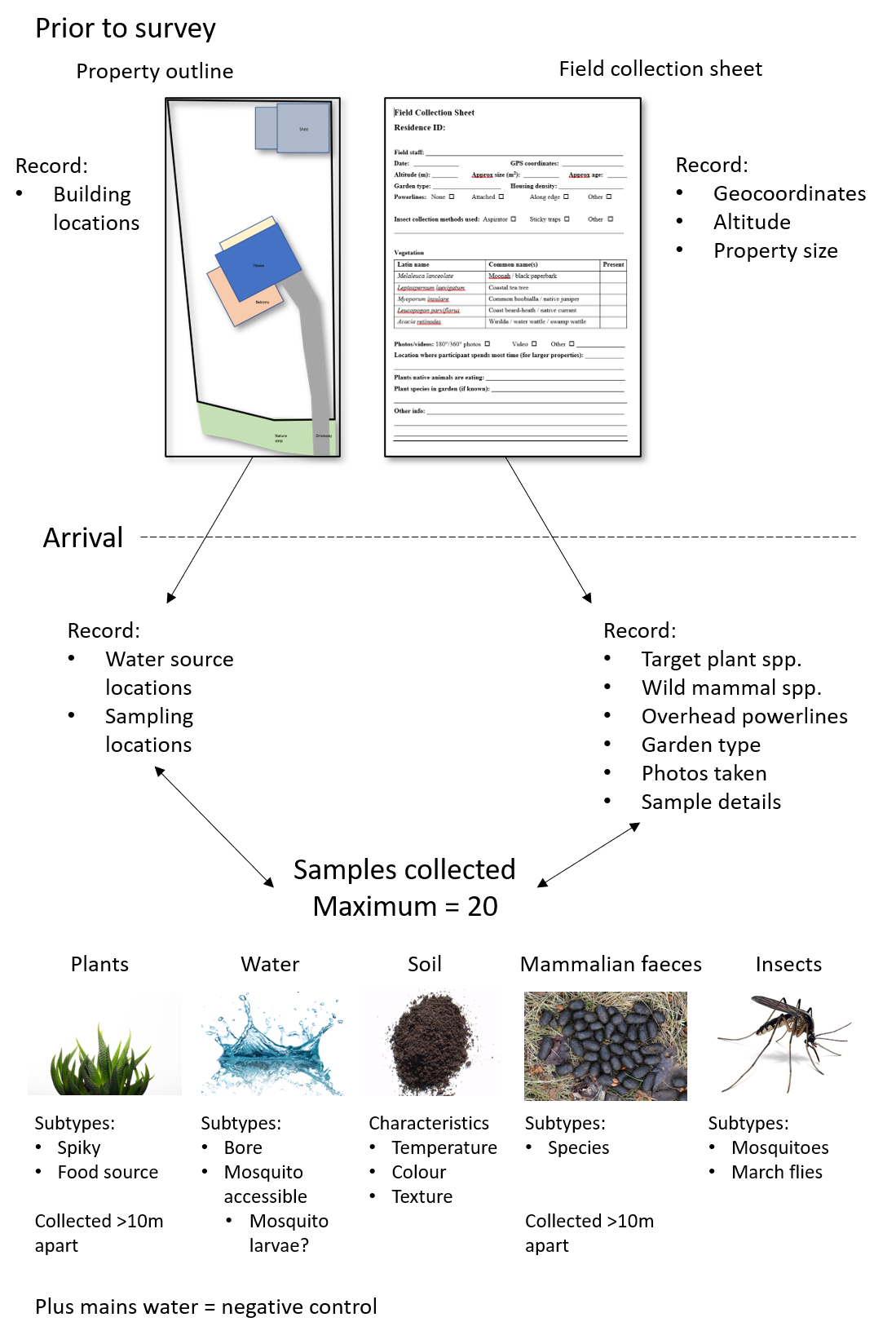

### Supplemental Figure 3

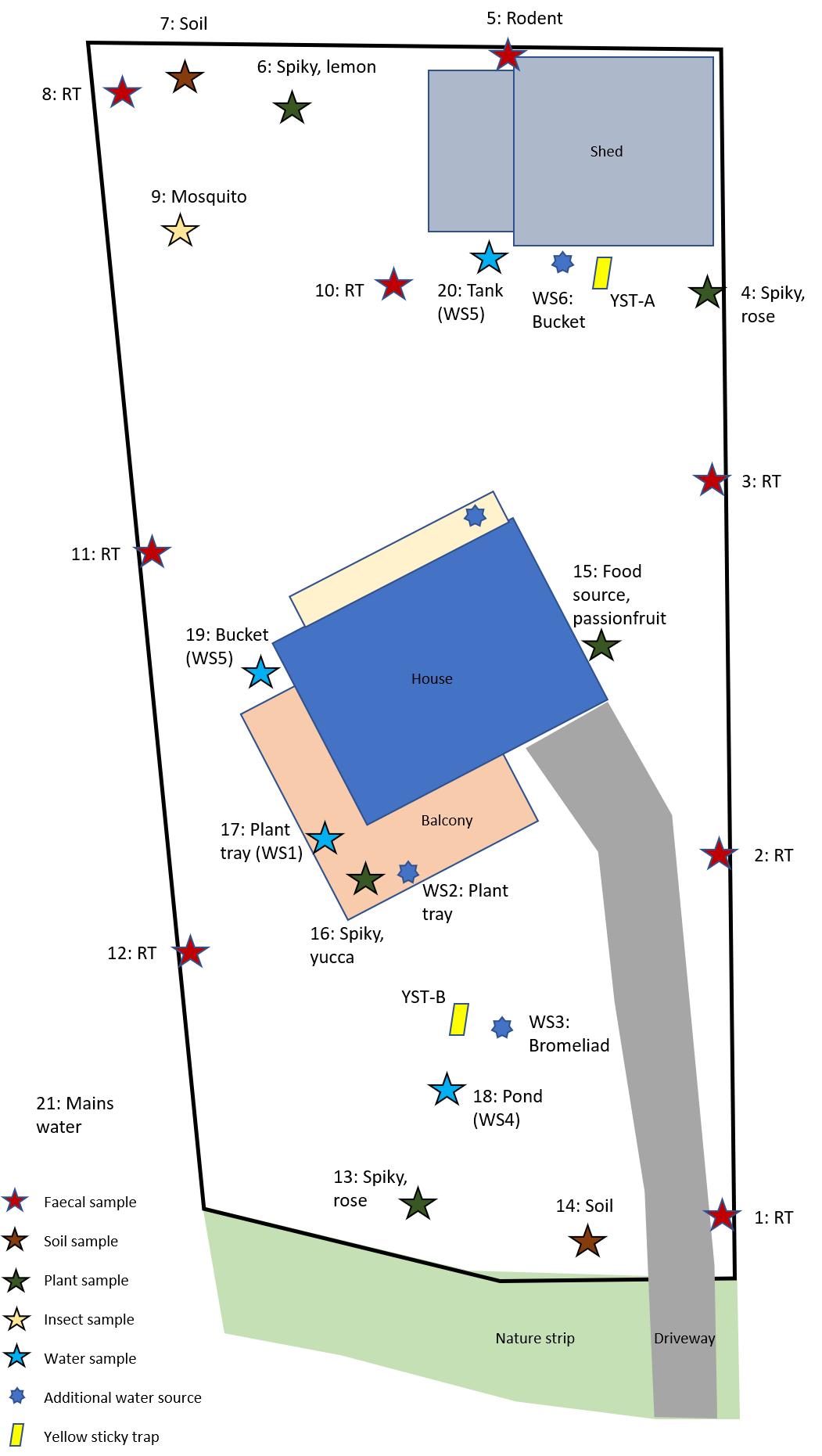

### Supplemental Figure 4

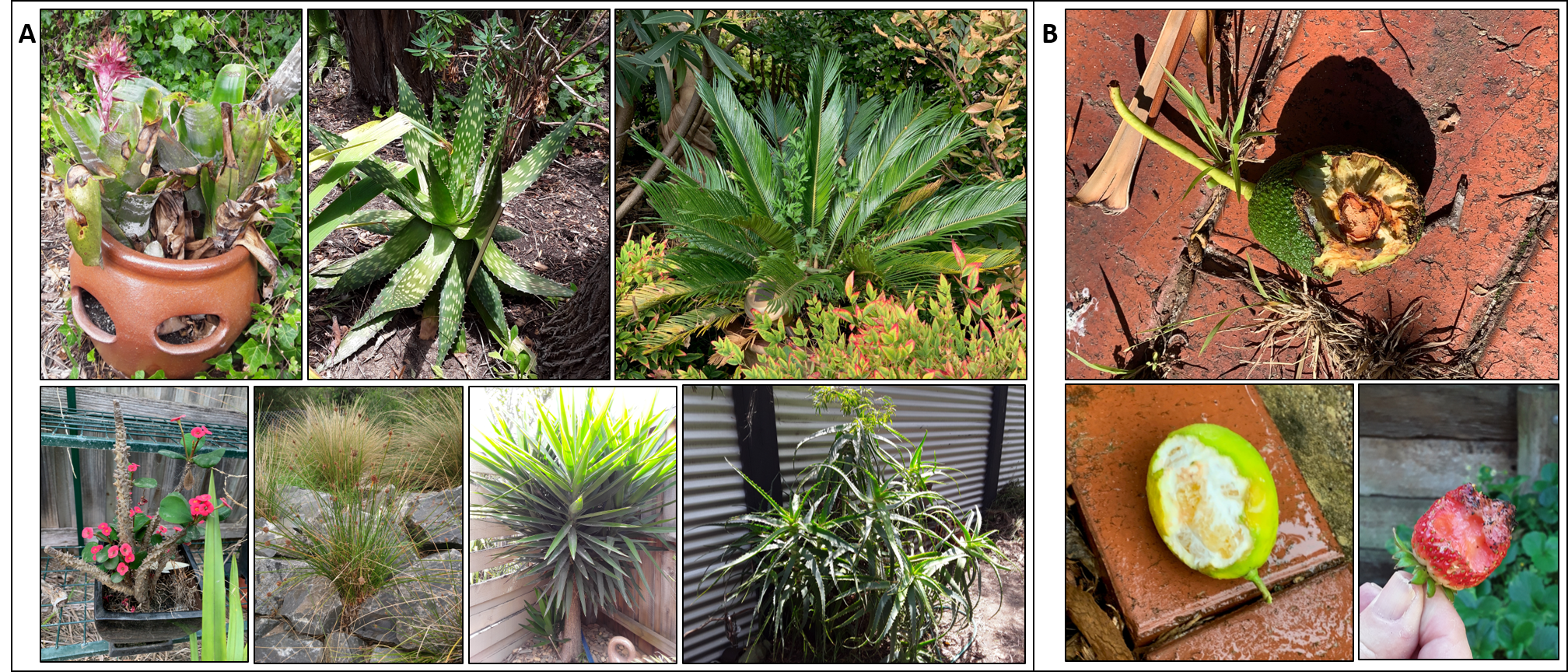

### Supplemental Figure 5

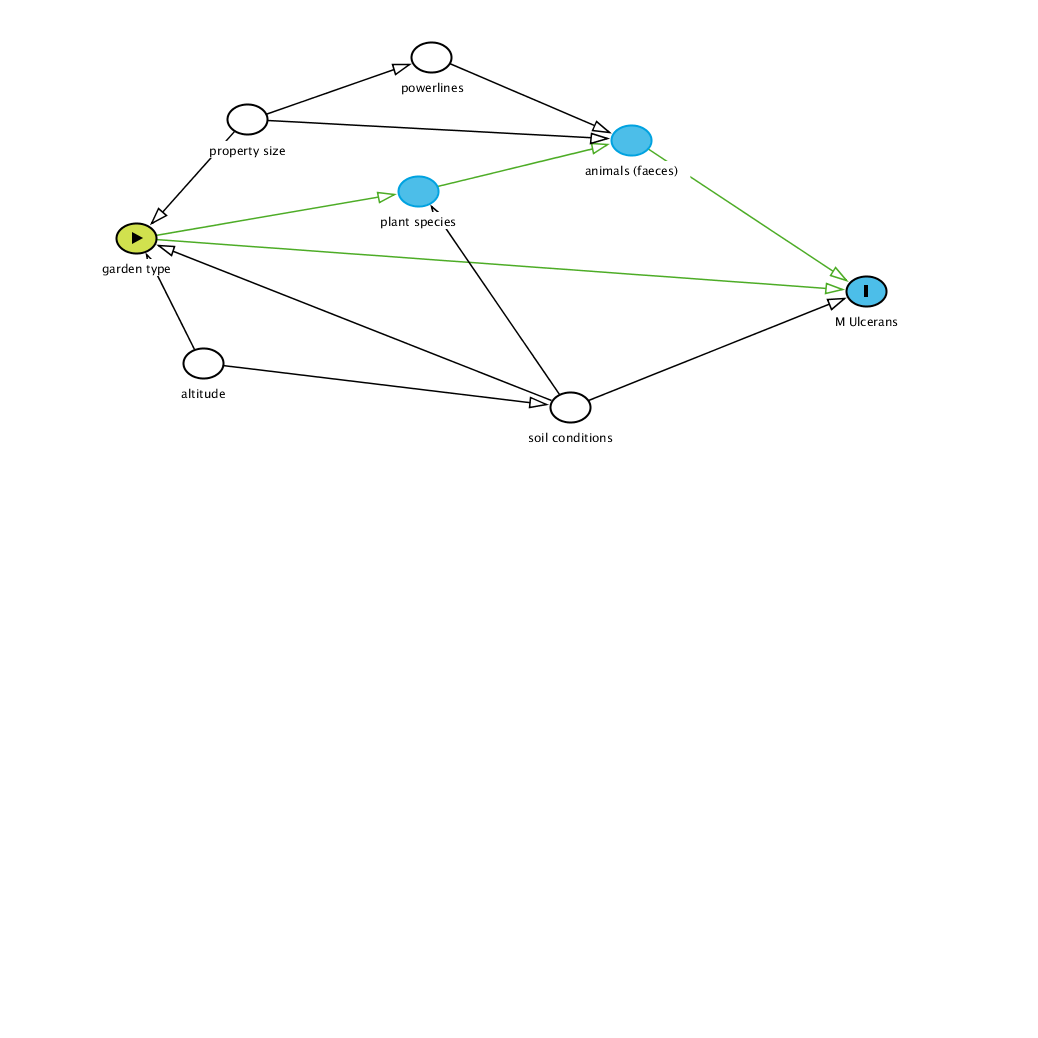

### Supplemental Figure 6

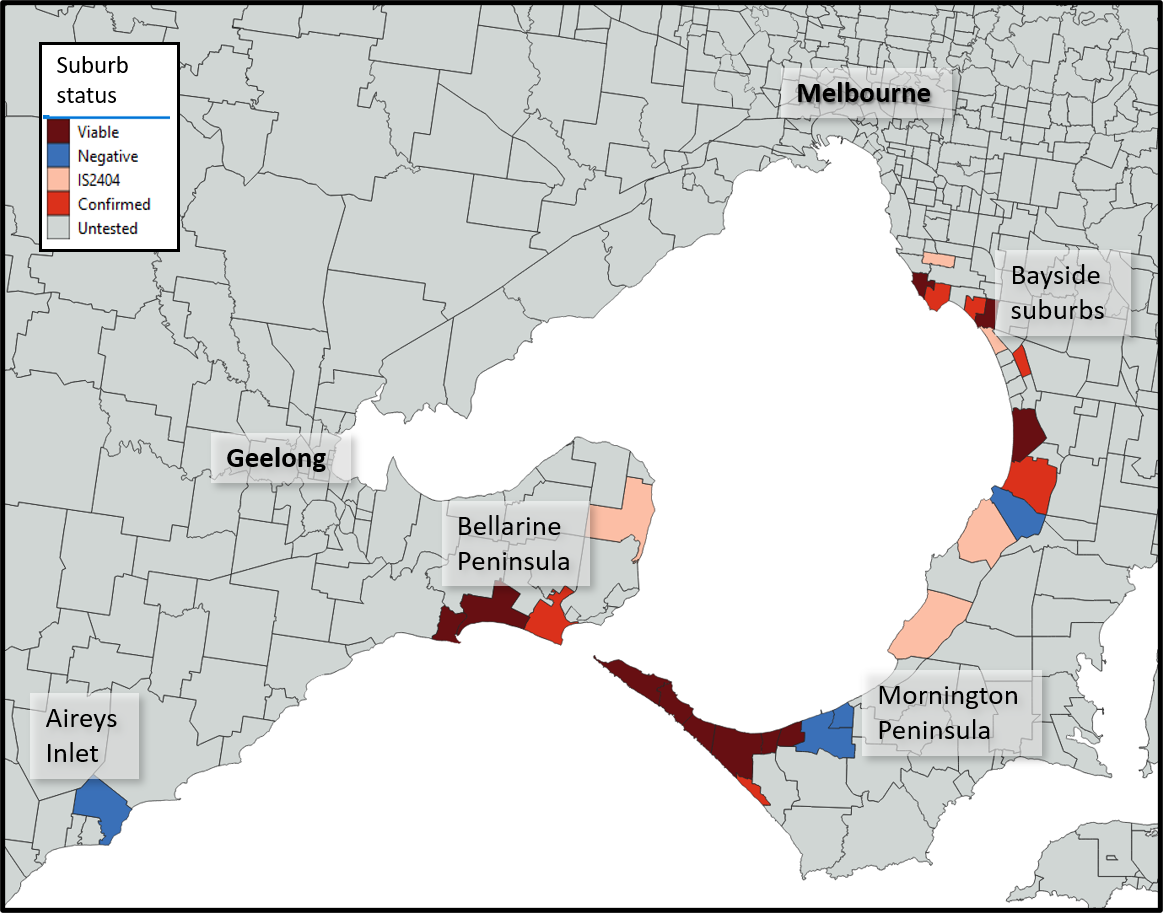
